# Case-level artificial intelligence for multi-photo teledermatology submissions: development and internal validation using patient-submitted dermatology images

**DOI:** 10.64898/2026.05.21.26353816

**Authors:** Vatsal Pravinbhai Patel, Nishi Seth, Abhijeet Patel, Yash Jayeshbhai Patel

## Abstract

**Background:** Store-and-forward teledermatology commonly relies on several patient-submitted photographs of the same concern, but most dermatology artificial intelligence models classify single images independently.

**Objective:** To develop and internally validate a case-level diagnostic-support model that aggregates multiple patient-submitted photographs for common dermatologic conditions.

**Methods:** We conducted a retrospective diagnostic-modeling study using the Skin Condition Image Network, a public dataset of deidentified self-taken dermatology images from US adults. We curated 2,336 cases comprising 5,041 images across 10 common inflammatory, allergic, and infectious conditions. Cases were split at the submission level into training, validation, and held-out test sets. Frozen general-purpose and dermatology-specific encoders were compared with image-level classifiers and a gated-attention multiple instance learning model that generated one case-level output from 1–3 images.

**Results:** The strongest image-level baseline, dermatology-specific embeddings with random forest classification, achieved macro/micro ROCAUCs of 0.797/0.854. Case-level aggregation improved discrimination, with dermatology-specific embeddings plus multiple instance learning achieving mean macro/micro ROC-AUCs of 0.819/0.863 across repeated stratified experiments. The locked final model achieved macro/micro ROCAUCs of 0.800/0.849 on the held-out test set. Balanced-threshold sensitivity/specificity examples were 0.702/0.688 for eczema and 0.818/0.826 for urticaria.

**Limitations:** Internal validation used a 10-condition subset from a US volunteer dataset; external validation, calibration, subgroup performance analysis, and prospective workflow studies are required.

**Conclusion:** Modeling the teledermatology submission as a multi-image case better reflects asynchronous dermatology workflow than single-image classification. The model is preliminary clinician-facing support for structured review and triage, not autonomous diagnosis.

**Key Points:**

- Store-and-forward teledermatology submissions usually contain multiple patient-submitted photographs, whereas most dermatology AI models classify single images independently.
- This study developed a case-level multiple instance learning model that aggregates 1–3 photographs from the same SCIN submission and produces one clinician-facing diagnostic-support output.
- Case-level aggregation modestly improved discrimination over the strongest image-level baseline and produced threshold-specific sensitivity/specificity outputs suitable for structured review and triage research.

## 1. Introduction

Skin disease is common, visually heterogeneous, and frequently first evaluated outside specialist dermatology settings. Primary care clinicians and teledermatology services are often asked to triage inflammatory, allergic, and infectious eruptions that overlap in morphology and may require different urgency, counseling, or referral pathways.^[1,2]^ For clinical dermatology, artificial intelligence (AI) is therefore most useful when it supports the workflow of human clinicians rather than attempting to replace clinical assessment.

Store-and-forward teledermatology is a particularly relevant setting for such support. In routine asynchronous care, patients or referring clinicians rarely submit a single idealized image. They more often submit several photographs of the same concern, such as a close-up view, an angled view, and a wider contextual image. The dermatologist’s task is case-level interpretation: judging whether the submitted material is adequate, generating a differential diagnosis, deciding whether more information is needed, and determining whether the patient requires routine, expedited, or in-person review.^[3–5]^

Most image-based dermatology AI studies do not fully match this workflow. Many models classify one image at a time, even though clinical reasoning in teledermatology often depends on the relationship among multiple views. A close-up image may show scale, vesiculation, follicular involvement, or crust. A contextual image may show distribution, symmetry, body-site pattern, or extent. Treating these images independently can discard information that clinicians naturally integrate during case review.

Recent dermatology AI research has shown both promise and caution. Deep learning systems can support differential diagnosis, and clinician–AI collaboration may improve diagnostic accuracy in store-and-forward simulations.^[1,6]^ However, studies also show that performance can vary across skin tones, disease groups, image quality, and clinical context, making intended use, transparency, subgroup evaluation, and cautious deployment essential.^[7,8]^ In clinical dermatology journals, the strongest AI papers now emphasize not only model performance but also clinical role, threshold interpretation, and limitations relevant to patient management.^[9,10]^

The Skin Condition Image Network (SCIN) provides a useful substrate for studying this problem because it contains deidentified, self-taken dermatology photographs contributed by US adults, with 1–3 images per submission, symptom and demographic information, dermatologist differential labels, and estimated Fitzpatrick Skin Type and Monk Skin Tone annotations.^[11]^ SCIN is especially relevant to teledermatology because most diagnosable submissions represent common allergic, infectious, or inflammatory concerns rather than the malignancy-heavy case mix of many historical dermatology AI datasets. At the same time, SCIN labels are retrospective dermatologist differentials rather than definitive encounter-confirmed diagnoses; models trained on this resource should therefore be interpreted as diagnostic-support systems, not autonomous diagnostic devices.

A case-level model requires a method that can aggregate a variable set of photographs into one submission-level output. Multiple instance learning (MIL) provides this formulation: a case is represented as a bag of image instances with one case-level label. Attention-based MIL further allows the model to learn which image views contribute most to the case-level prediction while preserving permutation invariance.^[12,13]^ Dermatology-specific foundation encoders also make this approach computationally practical by providing fixed image embeddings that can be paired with a compact downstream model rather than requiring full end-to-end retraining.^[14]^

We therefore developed and internally validated DermAssist, a case-level AI model for multi-photo teledermatology submissions. The model aggregates 1–3 patient-submitted photographs and produces case-level diagnostic-support probabilities across 10 common dermatologic conditions. We compared general-purpose and dermatology-specific frozen encoders, image-level baselines, and a gated-attention MIL model under matched case-wise splits. We evaluated discrimination using macro/micro ROC-AUC and precision-recall metrics, and we translated model outputs into clinically interpretable operating points using validation-selected thresholds, sensitivity, and specificity. The intended role is clinician-facing structured review and triage support in teledermatology or primary care workflows, not autonomous diagnosis.

## 2. Methods

### 2.1. Data

#### Primary source

We curated a case-level dataset from the Skin Condition Image Network (SCIN), an open-access collection of self-taken dermatology photographs contributed by US adults after online informed consent.^[11]^ Each submission contains 1–3 images, typically close-up, angled, and contextual views, together with optional demographic and symptom information. Images underwent third-party de-identification and quality screening in the source study. Each diagnosable case was labeled retrospectively by board-certified dermatologists who provided a differential diagnosis with confidence from a controlled dermatology vocabulary. The public release includes estimated Fitzpatrick Skin Type (eFST) and estimated Monk Skin Tone (eMST) annotations. Because these labels represent dermatologist differentials rather than definitive encounter-confirmed diagnoses, the present model is evaluated as diagnostic support rather than autonomous diagnosis.

#### Working cohort (10 conditions)

From SCIN’s broader taxonomy we construct a *10-condition* working set targeting common primary-care presentations: *Eczema, Allergic Contact Dermatitis, Insect Bite, Urticaria, Psoriasis, Folliculitis, Irritant Contact Dermatitis, Tinea, Herpes Zoster,* and *Drug Rash*. The resulting cohort comprises **2,336** cases (bags) and **5,041** images/embeddings referencing **5,032** unique image hashes. We preserve the natural multi-label structure (co-occurring diagnoses are common in dermatology).

#### Preprocessing and label construction

Starting from the public SCIN CSVs and embedding dumps, we apply the following steps before modeling:

1. *Eligibility and reliability.* Remove submissions marked non-diagnosable by SCIN reviewers or missing differentials; drop images missing on disk.
2. *De-duplication and cross-case conflicts.* Deduplicate image hashes and exclude rare hashes appearing under multiple case IDs with inconsistent labels (manifested in audit ledgers).
3. *Multi-label binarization.* Aggregate each case’s dermatologist differential into a *10-way multi-hot* vector: any non-zero weight for a kept class is treated as positive, all other kept classes negative. This preserves multi-diagnosis cases while restricting evaluation to the 10 targets.

**Figure 1:**
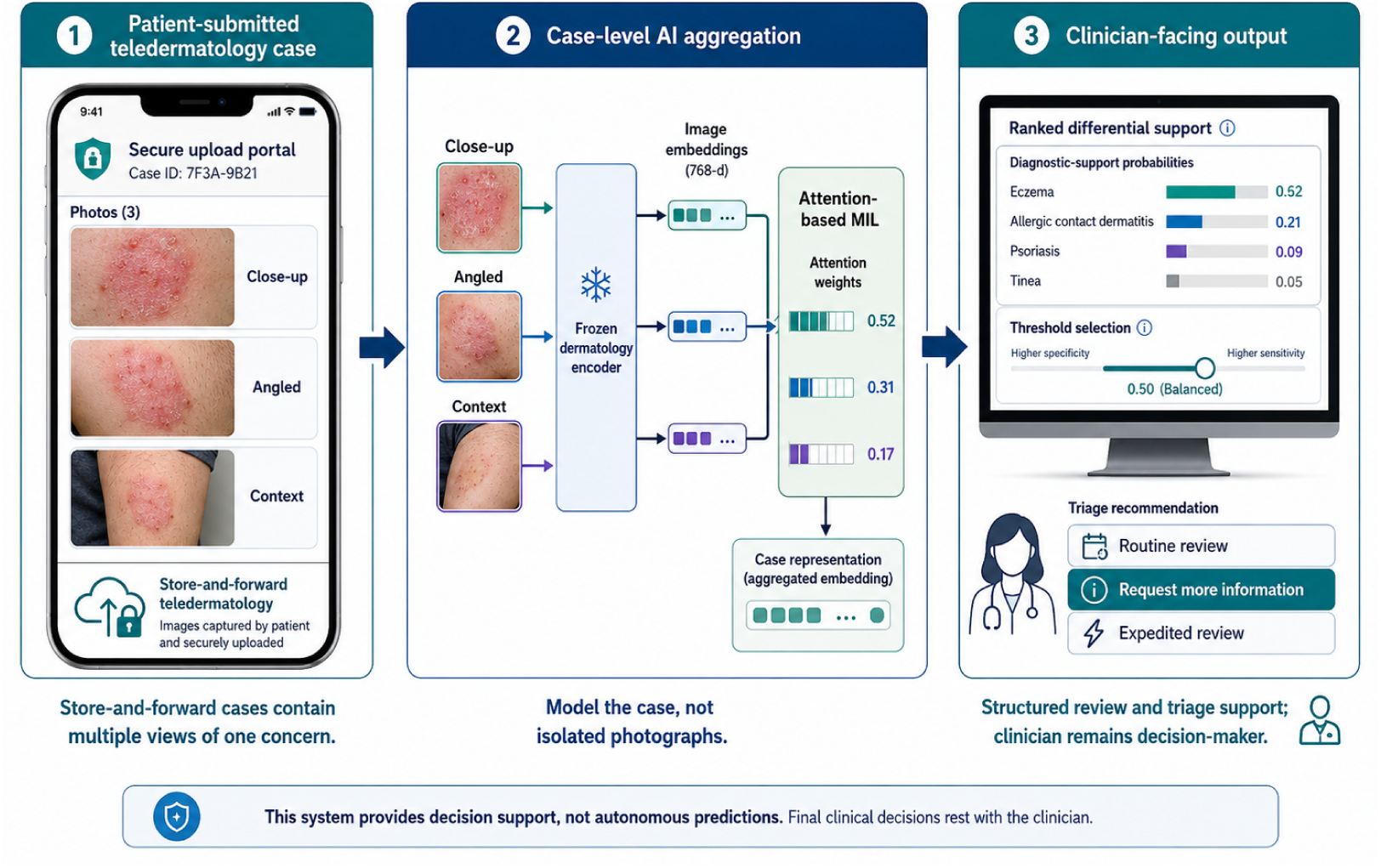
Case-level AI workflow for multi-photo teledermatology. Patient-submitted store-and-forward cases contain multiple views of one concern; DermAssist aggregates image embeddings with attention-based multiple instance learning and returns clinician-facing diagnostic-support and triage outputs.

#### Bag construction for MIL

For each case we select up to **three** images (prioritizing SCIN’s canonical triplet: close-up → angled → context). When more than three images exist, we keep the three best by SCIN quality indicators; when fewer are available, bags are padded to length three with masked placeholders to maintain permutation-invariant pooling. Thus each training example is a set *B_i_* = {*x_i_*_1_*, x_i_*_2_*, x_i_*_3_} with a multi-label vector *y_i_* ∈ {0, 1}^10^.

#### Summary statistics

Across the full cohort, the *median* number of images per case is **2** (IQR 1–3). The *mean* images per case are 2.16 (train), 2.13 (validation), and 2.18 (test). Approximately half of cases are multi-label after binarization. A complete per-class count table (cases and images across splits) is provided in the Results.

**Figure 2:**
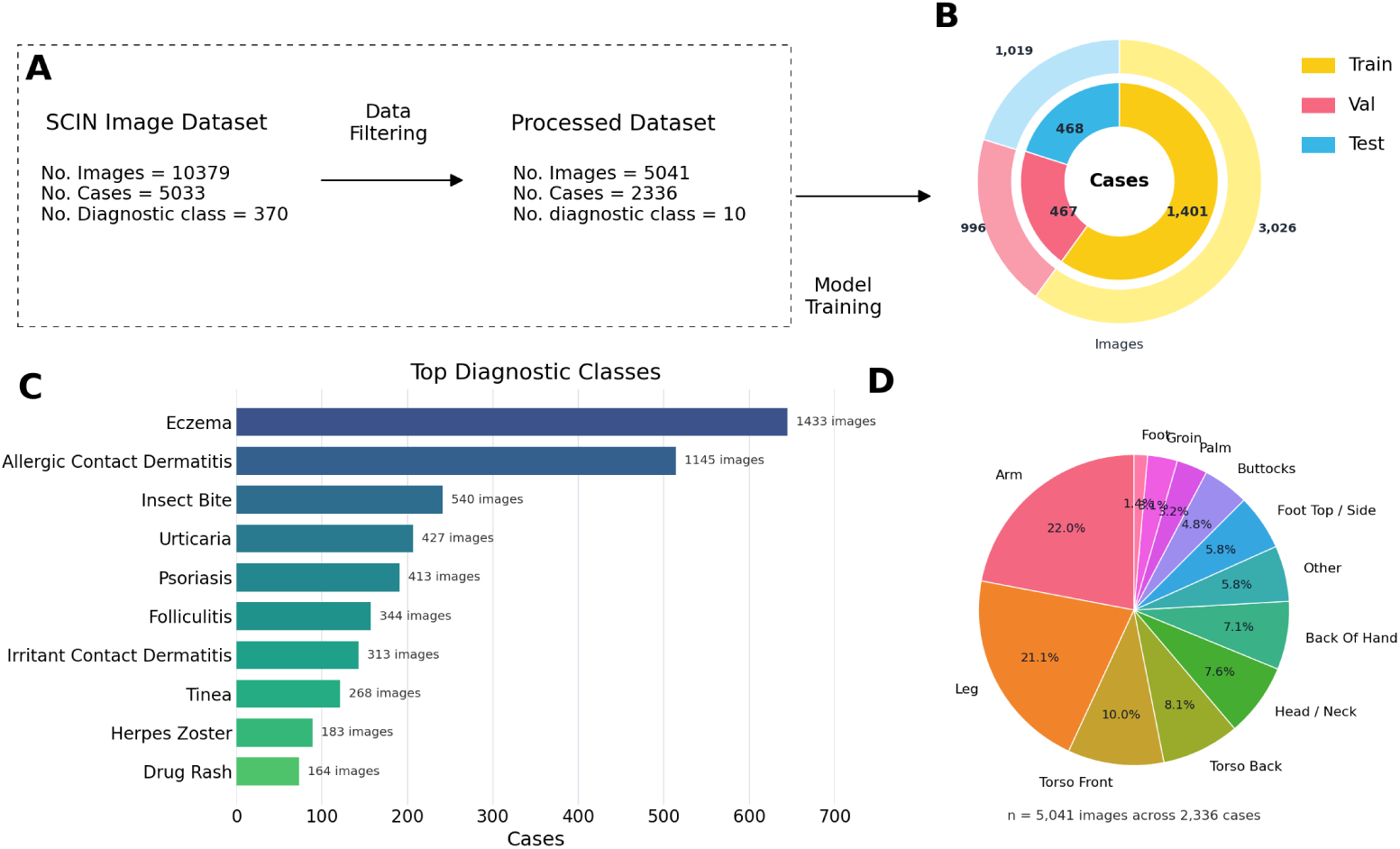
SCIN-derived teledermatology cohort and case-level preprocessing. **(A)** Flow diagram showing curation from the public SCIN resource to the DermAssist analytic cohort of 2,336 cases, 5,041 image rows/embeddings, and 10 target diagnostic classes. **(B)** Case-wise train, validation, and held-out test split composition. **(C)** Training-set case distribution across the 10 target conditions: eczema, allergic contact dermatitis, insect bite, urticaria, psoriasis, folliculitis, irritant contact dermatitis, tinea, herpes zoster, and drug rash. **(D)** Anatomical-site distribution across submitted images, illustrating the heterogeneous body-site coverage typical of patient-submitted teledermatology photographs.

#### Splits and held-out test set

We retain a fixed **test set** of **468** cases for all experiments. The remaining cases are partitioned into **train** (**1,401**) and **validation** (**467**) sets. All comparative baselines and MIL models use exactly these splits. Cross-validation used elsewhere in the paper is performed *within* the training pool and does not touch the fixed test set.

#### Governance and access

SCIN is released publicly with a dedicated *SCIN Data Use License* that governs redistribution and downstream use of the images and labels; we adhere to the license and report only aggregate statistics here. Repository documentation summarizes known issues (e.g., a small number of duplicate hashes retained to match the paper’s analysis) and provides schema details for diagnoses, eFST, and eMST.

### 2.2. Intended clinical role and end users

DermAssist is evaluated as a clinician-facing diagnostic-support model for asynchronous teledermatology and primary care workflows. The intended users are clinicians reviewing patient-submitted dermatology photographs, including primary care clinicians, teledermatology triage teams, and dermatologists managing asynchronous image queues. The model is designed to provide a ranked probability profile across a fixed 10-condition vocabulary and to support structured review, triage prioritization, and differential diagnosis generation. It is not intended to replace dermatologists, independently establish a diagnosis, determine treatment, or discharge patients without clinician review.

### 2.3. Training/validation/test preparation

We adopt a case-wise split to prevent any cross-set leakage of patient encounters or images. The DermAssist analytic cohort (10 diagnostic conditions) is divided 60/20/20 into train/validation/test at the case level using iterative stratification to preserve marginal label frequencies in a multi-label setting (order-1 label preservation). This procedure is widely used for multilabel tasks because it maintains per-class prevalence more faithfully than naive random or single-label stratification.

Table 2 summarizes counts per split. Image totals refer to rows in the embedding manifest; because nine image hashes appear in two distinct cases, the cohort contains 5,032 unique image hashes. Class-wise counts (cases and images) are visualized in *Top Diagnostic Classes* and the split composition in *MIL Dataset Composition* (see provided figures).

**Table 1:** Clinical interpretation of the SCIN source data and DermAssist analytic cohort.

| Item | Clinical interpretation |
| --- | --- |
| Source population | US adults who contributed self-taken dermatology photographs after online informed consent through the SCIN web application. |
| Case structure | Each submission contains 1–3 photographs of the same concern, typically including close-up, angled, and contextual views, matching the structure of many store-and-forward teledermatology cases. |
| Reference labels | Dermatologist-provided differential diagnoses, not biopsy-confirmed or encounter-confirmed final diagnoses. Model outputs should therefore be interpreted as diagnostic-support probabilities. |
| Skin-tone annotations | SCIN provides estimated Fitzpatrick Skin Type and estimated Monk Skin Tone labels, enabling future subgroup performance and fairness analyses. |
| Analytic cohort | 2,336 cases and 5,041 image rows/embeddings across 10 common inflammatory, allergic, and infectious conditions. |
| Unit of inference | The patient-submitted case, not the individual image. |
| Intended clinical role | Clinician-facing structured review and triage support for teledermatology or primary care; not autonomous diagnosis or treatment selection. |

**Table 2:** Cohort sizes by split using case-wise 60/20/20 partitioning. Image counts refer to image rows/embeddings; the cohort contains 5,032 unique image hashes because nine hashes appear in two distinct cases.

| Split | Cases | Images / embeddings |
| --- | --- | --- |
| Train | 1,401 | 3,026 |
| Validation | 467 | 996 |
| Test | 468 | 1,019 |
| Total | 2,336 | 5,041 |

For image-level baselines (Sec. 2.3), each image is treated as an independent instance but inherits the case’s multi-hot label vector. Crucially, every image from a case remains in that case’s assigned split, ensuring that the same visual content (or near-duplicates) never appears in both training and evaluation sets. For MIL experiments, each case contributes a bag containing up to three images (close-up, angled, context); bags with fewer views are padded and masked, preserving permutation invariance while enabling consistent batching.

Where indicated, cross-validation sweeps are performed within the training pool only; the validation split is used for model selection, early stopping, threshold calibration, and probability calibration, and the test split is held out for final reporting. Given the class imbalance typical of dermatology diagnosis, we report both ROC-AUC and PR-AUC, with PR-AUC emphasized in selection criteria.

### 2.4. Feature extraction (frozen encoders)

In all experiments we use frozen encoders to compute a fixed-length vector for each image; these embeddings are cached and serve as inputs to either image-level classifiers or bag-level MIL models. “Feature extraction” thus refers to running a pretrained vision backbone forward once per image (no back-propagation through the encoder) and retaining the penultimate or projection features as the representation used downstream.

We evaluate six encoders spanning classical CNNs, general-purpose transformers, and a dermatology-specific foundation model. Torch-based CNNs rely on canonical torchvision preprocessors, and transformer encoders on HuggingFace AutoImageProcessor, ensuring that resizing, normalization, and color space match pretraining. The Derm Foundation model is used via its public release, which provides 6,144-dimensional dermatology embeddings.

ResNet-50 and Inception-v3 provide strong convolutional baselines with efficient spatial inductive bias. BiT-50 tests large-scale supervised pretraining transfer. ViT-Base probes patch-wise self-attention without convolutions. CLIP ViT-B/32 evaluates language-aligned representations. Derm Foundation supplies domain-specific features explicitly optimized for skin images; using its released 6,144-D embeddings enables data- and compute-efficient downstream learning.

**Table 3:** Frozen encoders used for feature extraction. Parameter counts are approximate for standard published variants; embedding dimensionality reflects the vector used in this study.

| Encoder | Architectural note | Params (M) | Embedding | Preprocessor |
| --- | --- | --- | --- | --- |
| ResNet-50 | Residual bottleneck CNN<sup>[15]</sup> | ~25.6 | 2048 | torchvision |
| Inception-v3 | Factorized conv + aux head<sup>[16]</sup> | ~27.2 | 2048 | torchvision |
| BiT-50 | ResNet-50 pre-trained at scale<sup>[17]</sup> | ~23.5 | 2048 | torchvision |
| ViT-Base | 16×16 patch transformer<sup>[18]</sup> | ~86.4 | 768 | HF AutoImageProcessor |
| CLIP ViT-B/32 | Contrastive image-text transformer<sup>[19]</sup> | ~87.9 | 512 | HF CLIP processor |
| Derm Foundation | Domain-specific dermatology encoder | ~382 | **6144** | release-provided |

### 2.5. Image-level baseline classifiers

To contextualize MIL, we train per-image multi-label baselines on the frozen embeddings from each encoder. After fitting a StandardScaler on the training set features, we evaluate five families of classifiers chosen for complementary bias–variance profiles and widespread use in clinical ML:

- Logistic regression (one-vs-rest, class-weighted) for a calibrated linear baseline.
- Linear SVM with Platt (sigmoid) calibration via CalibratedClassifierCV, yielding probabilistic outputs suitable for threshold tuning.
- Random forest (300 trees, class-weighted) for non-linear ensembles robust to heterogeneous features.
- Gradient boosting trees for additive, stage-wise non-linear decision boundaries.
- k-nearest neighbours (k=5, distance weighting) as a non-parametric baseline.

All models produce per-class probabilities on validation and test splits. Thresholds for operating-point metrics are selected on the validation set (per class), while selection among candidates emphasizes macro/micro ROC-AUC with PR-AUC reported due to class imbalance. A heatmap summarizing test macro AUC across (encoder × classifier) combinations is provided (see *Test Macro ROC-AUC by Encoder and Classifier*).

### 2.6. Attention-based multiple-instance classifier

Each clinical encounter is modeled as a bag *B_i_*= {*x_i_*_1_*, x_i_*_2_*, x_i_*_3_} of up to three image embeddings with a multi-hot label vector *y_i_* ∈ {0, 1}^10^. We adopt the gated attention MIL operator of Ilse *et al.* to produce a permutation-invariant bag representation while attributing importance to individual views (close-up / angled / context). Concretely, each instance *x_ij_* is first projected by an MLP to *h_ij_* ∈ R*^d^* (ReLU activations). Attention scores are computed via parallel tanh and *σ* projections,

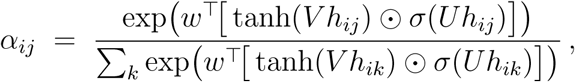

with a masked softmax to handle bags of length *<* 3. The bag embedding is *z_i_* = Σ*_j_α_ij_h_ij_*, which is passed through a small classifier head (Layer-Norm → MLP) to yield 10 sigmoid outputs. This architecture aligns with the case-level nature of dermatology encounters (sets of photos per complaint), handles missing views, and exposes interpretable per-image weights for qualitative review.

Training minimises a class-balanced binary cross-entropy (CB-BCE) objective to counter label skew: class weights follow the “effective number of samples” with parameter *β* ≈ 0.996, and the loss reduces to a weighted logistic objective without focal modulation. We optimise with AdamW to decouple weight decay (1.6 × 10^−6^) from the adaptive updates, use a learning rate of 1.1 × 10^−4^, omit dropout in the instance encoder, and trigger early stopping on validation ROC-AUC with patience five.

### 2.7. Hyperparameter search and training setup

The MIL head was tuned using Hyperband, a resource-aware early-stopping strategy that allocates more training budget to promising configurations. The search varied instance MLP widths, attention width, bag-head widths, dropout, optimizer settings, loss function, batch size, and thresholding scheme. The selected configuration used Derm Foundation embeddings, an instance MLP of (768, 128), attention width 128, bag-head (128, 64), classbalanced binary cross-entropy, AdamW optimization, learning rate 1.1×10^−4^, weight decay 1.6 × 10^−6^, batch size 16, and early stopping. Detailed search ranges and top Hyperband trials are reported in the Supplementary Methods and Supplementary Table E.6.

### 2.8. Cross-validation protocol and reporting

Model selection and uncertainty estimation use *K*=5 fold cross-validation performed *within* the training pool. Folds are created with iterative multi-label stratification to preserve marginal label frequencies (order-1) across splits, a method shown to better match per-class prevalence than naive random or single-label stratification. We fix seeds 125–129 for reproducibility and train each fold for up to five epochs under the selected configuration from Sec. 2.7. The validation fold drives early stopping and threshold tuning; the held-out test set remains untouched until final evaluation.

We report micro/macro ROC-AUC and PR-AUC (with PR-AUC emphasized for class imbalance), together with thresholded operating-point metrics derived on the validation split. Aggregated fold-level results (mean ± SD) for the Derm-Foundation + MIL setting are summarized in the accompanying figure (*Validation ROC-AUC per fold*) and in the machine-readable summary provided with the manuscript.

### 2.9. Class-imbalance handling and optimization

Because the 10 target conditions were imbalanced, all MIL experiments used class-balanced binary cross-entropy with effective-number reweighting computed from the training split only.^[20]^ This approach increases the loss contribution of rarer diagnoses while avoiding oversampling images across case-wise splits. Optimization used AdamW with early stopping on validation performance.^[21,22]^ Threshold selection was decoupled from model training: after training, per-class thresholds were selected on the validation split and then applied unchanged to the held-out test set. Mathematical details of the class-balanced loss are provided in the Supplementary Methods.

### 2.10. Operating-point selection

We re-score the Google Derm + DermMIL model across ten stratified bag seeds (seeds 0531–3593). For each diagnostic class the decision threshold *τ* is swept from 0.00 to 1.00 in 0.01 increments, computing case-level sensitivity TP*/*(TP + FN) and specificity TN*/*(TN + FP) per seed. The perseed traces are aligned on the common grid and averaged to obtain smooth mean±standard-deviation envelopes that capture cross-seed variability. The production operating point for a class is chosen as the smallest *τ* where the absolute difference |mean *Se* − mean *Sp*| is minimal; in the rare case of a tie we retain the lower *τ* to avoid artificially suppressing recall. This sweep also exposes auxiliary operating points (default 0.5, max-F1, Youden-J) that support the analyses in Section 3.

### 2.11. Evaluation metrics and clinical interpretation

The primary discrimination metrics were macro and micro receiver operating characteristic area under the curve (ROC-AUC). Macro ROC-AUC weights each diagnostic class equally and is useful when rare conditions should not be dominated by common conditions. Micro ROC-AUC pools all label decisions and therefore reflects overall discrimination across the cohort. Precision-recall AUC was also reported because positive labels are imbalanced across conditions.^[23]^

For clinical interpretability, we additionally report threshold-based sensitivity, specificity, precision, recall, F1 score, and balanced accuracy where available. ROC-AUC should be interpreted as a threshold-free ranking measure, not as the proportion of correct diagnoses. Sensitivity and specificity at validation-selected thresholds provide the more clinically intuitive operating characteristics for triage or structured review.

An exploratory comparison with closed multimodal assistants is described in Appendix Appendix A; this analysis was not used for model selection or primary interpretation.

## 3. Results

### 3.1. Cohort and teledermatology case structure

The analytic cohort included 2,336 patient-submitted cases and 5,041 image rows/embeddings from SCIN. The median number of images per case was 2 (IQR 1–3), reflecting the multi-photo structure of store-and-forward teledermatology submissions. The 10 target conditions represented common inflammatory, allergic, and infectious presentations: eczema, allergic contact dermatitis, insect bite, urticaria, psoriasis, folliculitis, irritant contact dermatitis, tinea, herpes zoster, and drug rash.

Cases were partitioned at the submission level into 1,401 training cases, 467 validation cases, and 468 held-out test cases (Table 2). This case-wise split ensured that images from the same submission did not appear in more than one partition. The cohort included all estimated Fitzpatrick Skin Types, although types II–III were most common. Because several condition-by-skintone strata were sparse, formal subgroup performance comparisons were not treated as definitive in this internal validation study.

### 3.2. Image-level baselines

Among image-level baselines, dermatology-specific embeddings provided the strongest feature representation. The best image-level model used Derm Foundation embeddings with random forest classification and achieved macro/micro ROC-AUCs of 0.797/0.854 on the held-out test set. General-purpose transformer encoders outperformed older convolutional baselines in several comparisons, but they did not exceed the dermatology-specific encoder under matched splits (Figure 3).

**Figure 3:**
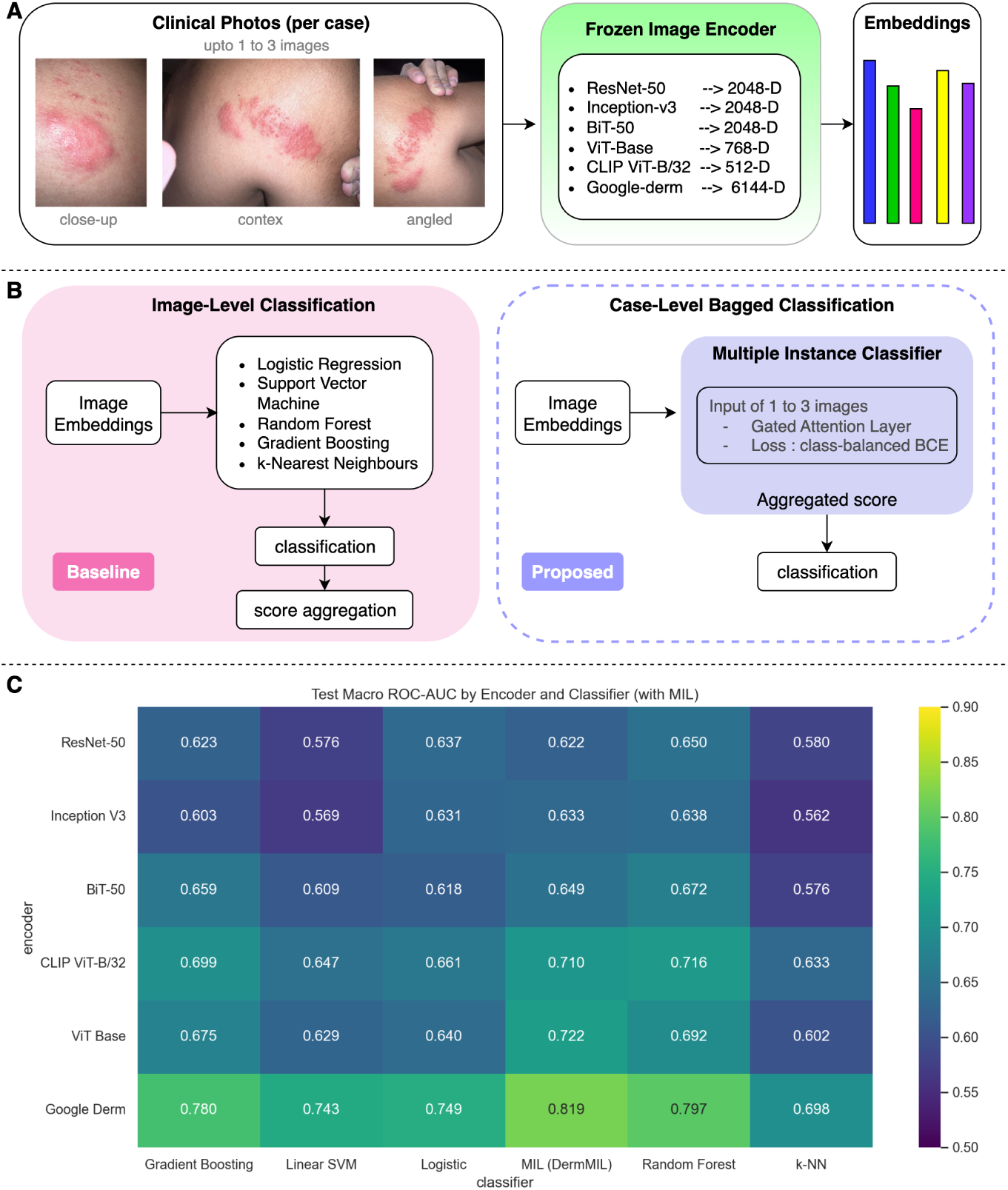
Pipeline and baseline performance across encoders. **(A)** Schematic of the DermAssist pipeline: each case comprises up to three clinical photos (close-up, angled, context). Features are extracted using frozen encoders (ResNet-50, Inception-v3, BiT-50, ViT-Base, CLIP ViT-B/32, Google Derm) producing 512–6,144D embeddings. **(B)** Comparison between image-level classification and case-level Multiple Instance Learning. Baselines aggregate independent image scores; DermMIL aggregates image embeddings via gated attention pooling to form a single case-level prediction. **(C)** Heatmap showing test macro ROC-AUCs across encoder-classifier pairs. Transformer and domain-specific encoders outperform CNNs, with Google Derm plus MIL achieving the highest mean macro ROC-AUC (0.819 across repeated stratified experiments).

### 3.3. Case-level multiple instance learning

The case-level MIL formulation improved discrimination over the strongest image-level baseline. Across 10 repeated stratified experiments, Derm Foundation embeddings plus gated-attention MIL achieved mean macro/micro ROC-AUCs of 0.819/0.863. The absolute macro ROC-AUC improvement over the best image-level baseline was 0.022. This improvement was modest but clinically relevant to the study question because the MIL model evaluates the submitted case as a multi-image unit rather than treating each photograph independently.

The locked final model achieved macro/micro ROC-AUCs of 0.800/0.849 on the held-out test set. These values indicate moderate case-level discrimination across a heterogeneous, multi-label, patient-submitted image cohort. Training curves and fold-level diagnostics are provided in the Supplementary Results because they support reproducibility but are less central to clinical interpretation.

### 3.4. Clinically interpretable operating points

At the default probability threshold of 0.5, the locked final model was highly conservative, producing high specificity but low sensitivity for several conditions. Validation-selected balanced thresholds produced more clinically interpretable tradeoffs. For example, eczema achieved sensitivity/specificity of 0.702/0.688 and urticaria achieved 0.818/0.826 at their respective balanced thresholds (Figure 4; Figure 5; Table 4). These operating points illustrate how the same model can be tuned toward higher-sensitivity screening or more balanced structured review, depending on the clinical workflow.

**Figure 4:**
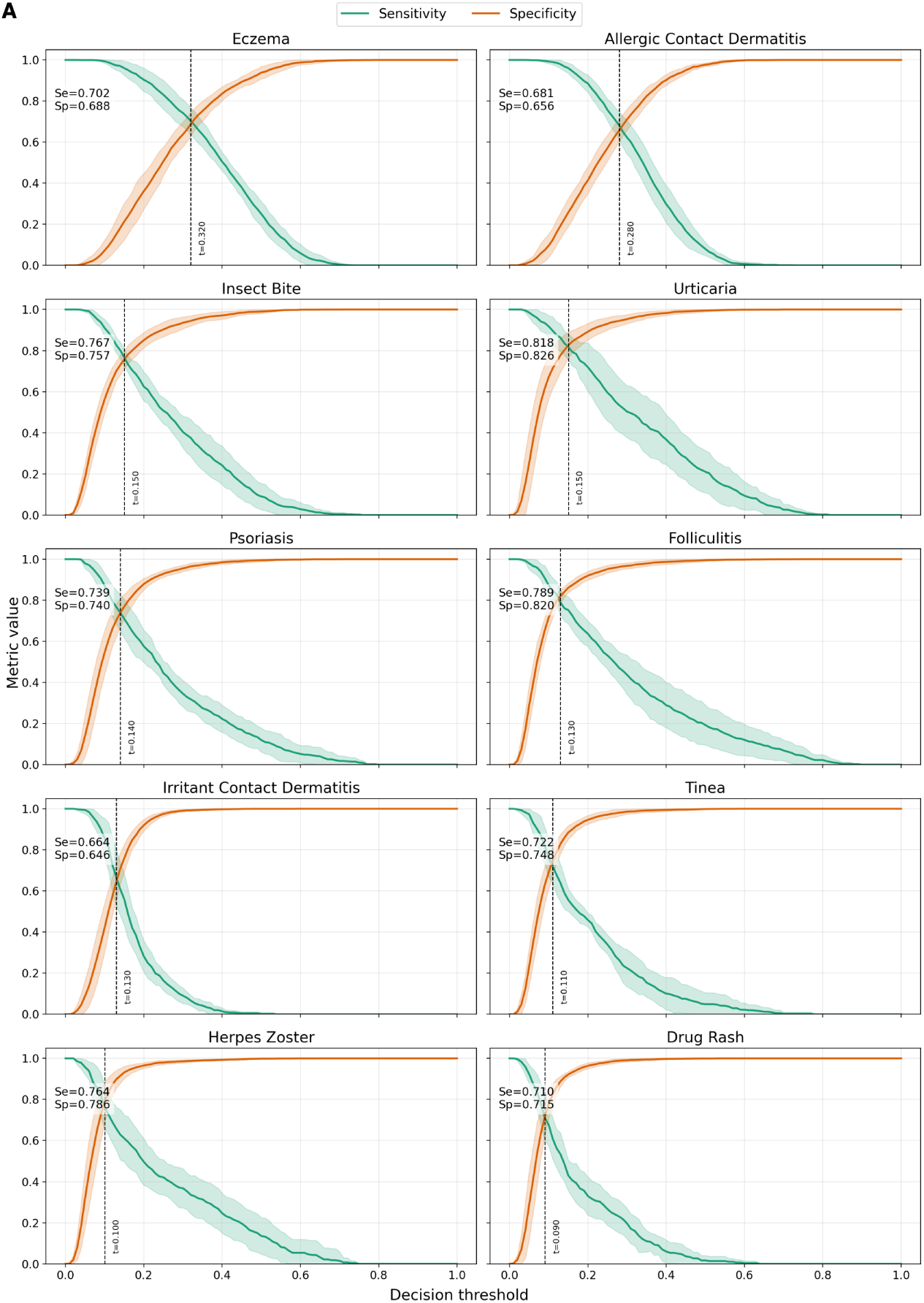
Per-condition threshold optimization. Each subplot depicts the relationship between sensitivity and specificity as a function of the decision threshold for one of the ten diagnostic conditions (10-run mean ± SD across stratified bag seeds). The dashed vertical line marks the balance-point threshold, defined as the smallest *τ* where the absolute difference between mean sensitivity and specificity is minimised. Reported sensitivity and specificity values correspond to that operating point.

**Figure 5:**
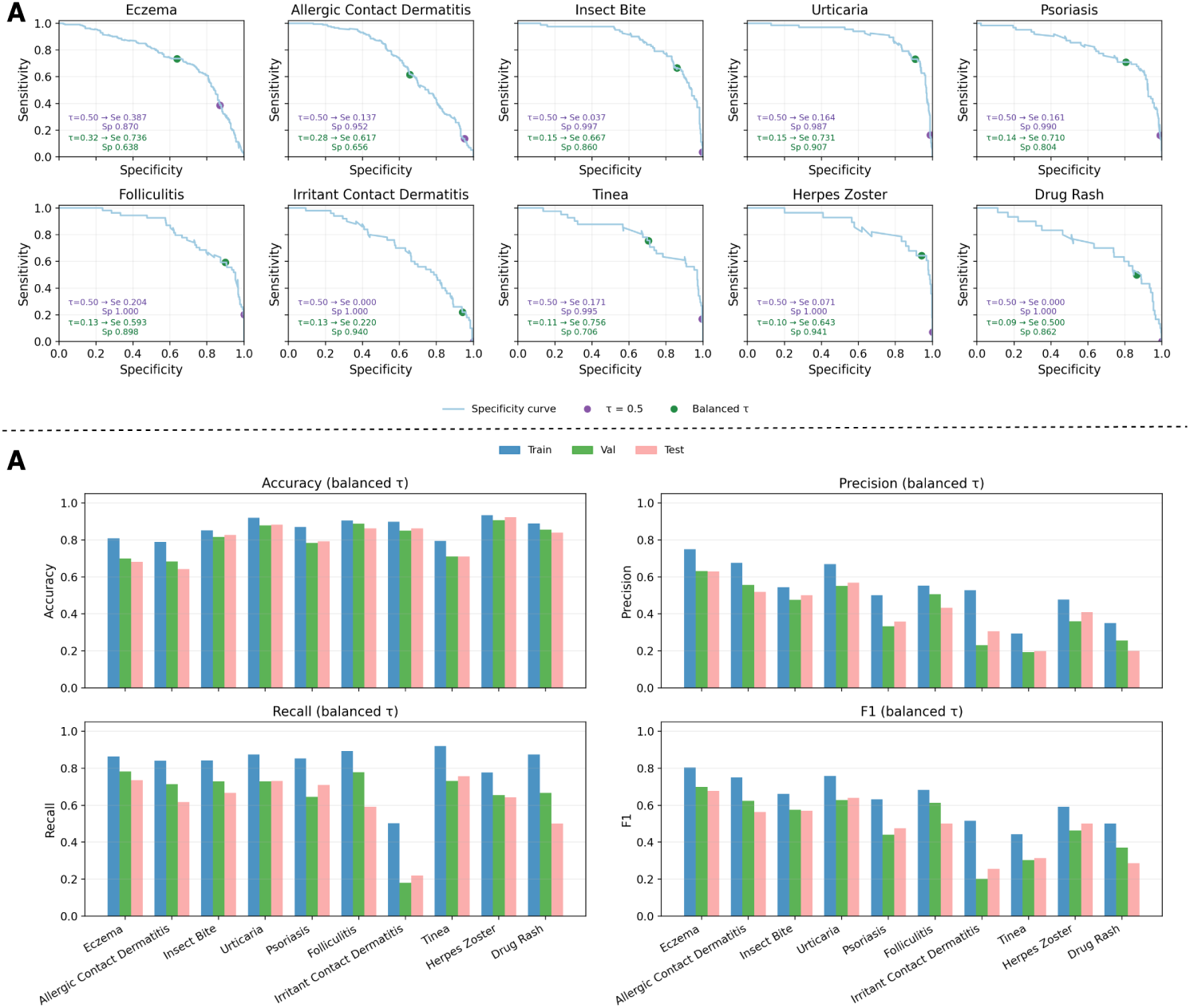
Clinician-readable operating-point performance. **(A)** Per-class accuracy, precision, recall, and F1 score on train, validation, and held-out test splits after applying validation-selected balanced thresholds. **(B)** Test-set sensitivity–specificity curves showing how threshold selection changes the clinical operating point for each condition. The default 0.5 threshold is conservative for several diagnoses; balanced thresholds move the model toward more clinically interpretable sensitivity/specificity tradeoffs.

**Table 4:**
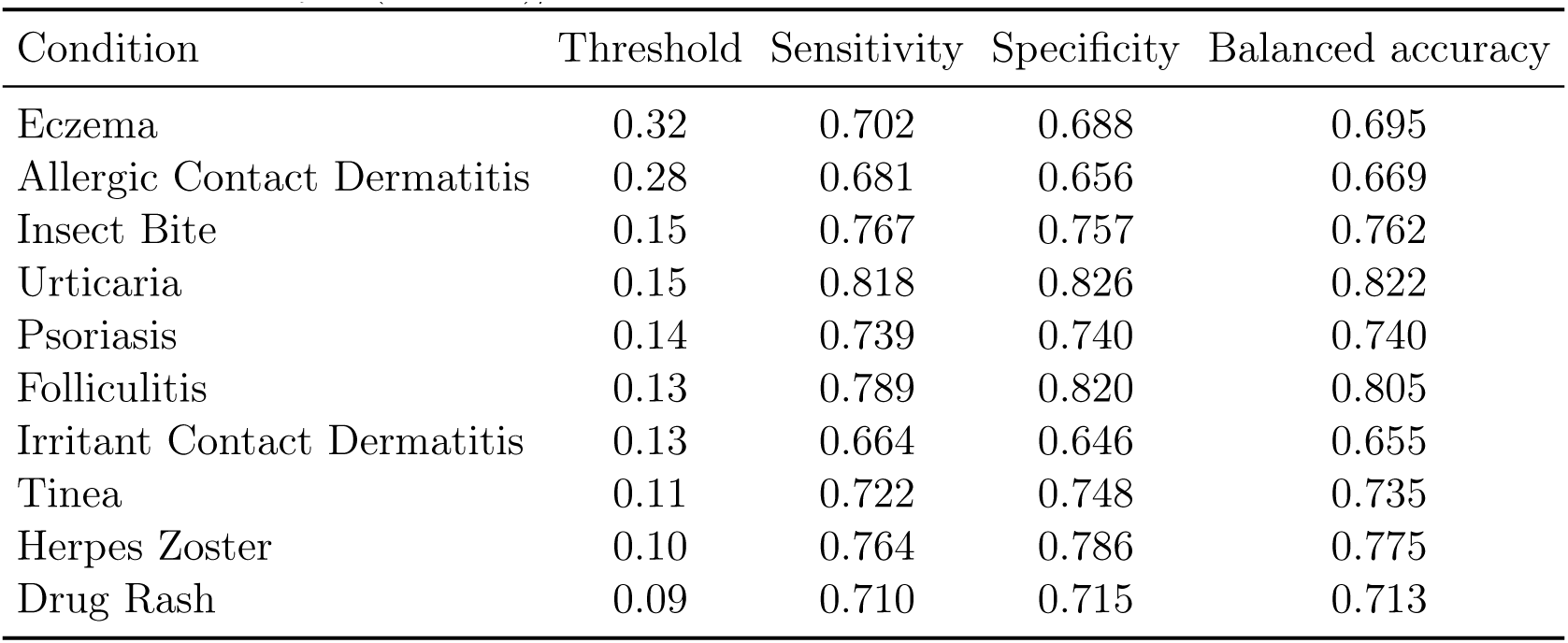
Balanced-threshold operating points for the 10 target conditions. Sensitivity and specificity are reported as 10-run means at the smallest threshold minimizing |*Se* − *Sp*|. Balanced accuracy is (*Se* + *Sp*)*/*2.

| Condition | Threshold | Sensitivity | Specificity | Balanced accuracy |
| --- | --- | --- | --- | --- |
| Eczema | 0.32 | 0.702 | 0.688 | 0.695 |
| Allergic Contact Dermatitis | 0.28 | 0.681 | 0.656 | 0.669 |
| Insect Bite | 0.15 | 0.767 | 0.757 | 0.762 |
| Urticaria | 0.15 | 0.818 | 0.826 | 0.822 |
| Psoriasis | 0.14 | 0.739 | 0.740 | 0.740 |
| Folliculitis | 0.13 | 0.789 | 0.820 | 0.805 |
| Irritant Contact Dermatitis | 0.13 | 0.664 | 0.646 | 0.655 |
| Tinea | 0.11 | 0.722 | 0.748 | 0.735 |
| Herpes Zoster | 0.10 | 0.764 | 0.786 | 0.775 |
| Drug Rash | 0.09 | 0.710 | 0.715 | 0.713 |

### 3.5. Exploratory comparison with general-purpose multimodal assistants

Exploratory zero-shot evaluation of proprietary multimodal assistants is reported in Appendix Appendix A. These results were not used as primary evidence because model versions, prompting behavior, and probability calibration can change over time. The primary finding of the present study remains the comparison between image-level classification and case-level MIL under matched, reproducible splits.

## 4. Discussion

In this retrospective diagnostic-modeling study, a case-level MIL model modestly improved discrimination over the strongest image-level baseline while better matching the structure of store-and-forward teledermatology submissions. The principal finding is not that the model is ready for autonomous diagnosis, but that modeling the submitted case as a set of photographs is a more clinically faithful formulation than classifying each image independently. This distinction matters because clinicians reviewing teledermatology cases integrate multiple views of the same concern, rather than treating each photograph as a separate patient encounter.

The study addresses a practical translational gap in dermatology AI. Many image-based models have been developed on single, curated images, often with an emphasis on neoplasms or dermoscopic lesions. By contrast, SCIN contains patient-submitted photographs of common conditions, and most diagnosable submissions represent inflammatory, allergic, or infectious concerns.^[11]^ This case mix is closer to the front-door problems encountered in primary care and asynchronous teledermatology, where clinicians often triage rashes, dermatitis-like eruptions, infections, and drug-related presentations from imperfect smartphone images.

The intended clinical role is structured review and triage support, not replacement of dermatologist judgment. A primary care clinician or teledermatology triage team might use a ranked probability profile to organize the differential diagnosis, identify cases requiring expedited review, or decide when additional images or history are needed. A dermatologist managing an asynchronous image queue might use the output as a second-reader signal. The AAD teledermatology standards emphasize clinician-directed care, access to board-certified dermatologists, the option for in-person care when needed, and adequate history and examination before prescribing; these principles are consistent with positioning DermAssist as supportive rather than autonomous.^[3]^

The threshold analysis is central to clinical interpretation. ROC-AUC summarizes discrimination across possible thresholds, but it does not tell a clinician how the model behaves at a specific operating point. At a default threshold of 0.5, the model was conservative for several diagnoses, with high specificity but low sensitivity. Balanced thresholds produced more clinically interpretable sensitivity/specificity tradeoffs. This presentation is deliberately closer to clinical diagnostic-test reporting than to a purely technical leaderboard. It also parallels the direction of recent clinically framed dermatology AI work, including AJCD studies that translate model outputs into severity or decision-support thresholds rather than reporting AUC alone.^[9,10]^ The modest size of the performance gain should be interpreted carefully.

The improvement from macro ROC-AUC 0.797 for the strongest image-level baseline to 0.819 for the repeated case-level MIL model suggests that multiphoto case structure contains useful information. However, this is not a large enough improvement to claim clinical effectiveness by itself. The value of the approach lies in its workflow alignment, computational efficiency, and interpretability: frozen dermatology embeddings reduce training burden, the MIL head aggregates 1–3 images into a case-level output, and attention weights can indicate which submitted view contributed most to the prediction. These properties make the model suitable for further clinical evaluation.

This work also fits into a broader movement toward assistive, workflow-aware AI in dermatology. Large reader studies show that AI can improve diagnostic accuracy in store-and-forward dermatology simulations, but also demonstrate that performance gaps across skin tones can persist or even widen depending on the user group and interface design.^[6]^ Reporting standards such as CLEAR Derm emphasize data transparency, technical assessment, intended use, and clinical application because image-based dermatology AI can fail when datasets, labels, or deployment context are poorly specified.^[7]^ Recent BJD discussions of AI in dermatology pathways similarly stress that clinical impact, governance, monitoring, and safety nets are necessary before AI can be responsibly integrated into care pathways.^[3,24,25]^ Several limitations are important. First, this was an internal validation study using a 10-condition subset of SCIN. The model has not been externally validated in a health-system teledermatology service, primary care network, or prospectively collected cohort. Second, SCIN is a US volunteer dataset recruited through web search advertisements, and the cohort does not necessarily represent all patients who seek dermatology care. Third, the reference labels are retrospective dermatologist differentials rather than final clinical diagnoses confirmed by longitudinal follow-up, biopsy, microbiology, or treatment response. Fourth, although SCIN provides eFST and eMST annotations, the current study did not establish definitive subgroup fairness or bias performance because several class-by-subgroup strata are sparse. Future work should report subgroup discrimination and calibration across skin tone, age, sex, body site, image quality, and image count per case. Fifth, probability calibration and decision-curve analysis were not fully established, and both are needed before clinical implementation.

The exploratory comparison with closed multimodal assistants should also be interpreted cautiously. General-purpose multimodal systems are appealing because they can process images and text, but zero-shot evaluations are sensitive to prompt design, model version, access date, output parsing, and calibration. For this reason, those results are kept supplementary and should not distract from the main scientific contribution: a reproducible, case-level model that directly tests whether multi-photo aggregation improves dermatology AI evaluation.

Future work should move from internal validation to clinical translation. The next steps are external validation on independent teledermatology datasets, prospective evaluation in clinician workflows, calibration analysis, subgroup performance reporting, and integration of structured clinical context such as age, body site, symptom duration, pain, pruritus, medication exposure, and immune status. The model should also be evaluated for its effect on referral quality, time-to-review, clinician workload, missed urgent diagnoses, and patient outcomes. These studies are necessary before considering deployment.

Overall, the findings support case-level modeling as a clinically appropriate direction for teledermatology AI. DermAssist shows that aggregating multiple patient-submitted photographs can modestly improve discrimination while providing threshold-specific outputs that clinicians can interpret. The model should be viewed as a preliminary structured-review aid for teledermatology research, not as a diagnostic device ready for independent clinical use.

## 5. Conclusion

DermAssist demonstrates that patient-submitted teledermatology cases can be modeled more faithfully as multi-image submissions than as isolated photographs. In this internal validation study, a dermatology-specific frozen encoder combined with gated-attention multiple instance learning produced modestly higher case-level discrimination than the strongest image-level baseline and yielded threshold-specific sensitivity/specificity outputs that are easier for clinicians to interpret.

The model is best positioned as preliminary clinician-facing support for structured review and triage in teledermatology or primary care workflows. External validation, calibration, subgroup performance analysis, and prospective workflow studies are required before clinical use. The broader implication is that dermatology AI should increasingly be evaluated at the level of the clinical encounter, not only the individual image.

## Acknowledgments

The authors have no additional acknowledgments to report.

## Additional Information

## Author Contributions

Conceptualization: VPP, YJP, AP. Methodology: VPP, YJP, AP. Software: VPP. Formal Analysis: VPP, YJP, NS, AP. Data Curation: VPP. Writing – Original Draft Preparation: VPP. Writing – Review and Editing: VPP, NS, AP, YJP. Supervision: VPP. All authors contributed to the article and approved the submitted version.

## Data Availability

The datasets analyzed during the current study are available in the Skin Condition Image Network (SCIN) repository, https://github.com/google-research-datasets/scin.

## Code Availability

The code for the DermAssist model and evaluation pipeline is available upon request from the corresponding author and will be open-sourced on GitHub following publication.

## Ethics Approval

This study was conducted using de-identified, publicly available data from the Skin Condition Image Network (SCIN). The SCIN project was approved by an independent Institutional Review Board (IRB), and all participants provided informed consent.

## Consent to Participate

Not applicable. No individual patient data was directly collected by the authors; all data was sourced from an existing, consented, and de-identified dataset.

## Consent for Publication

Not applicable. No individual patient data or identifying images are used in this manuscript.

## Abbreviations and Acronyms

AI: Artificial intelligence
AUC: Area under the curve
CLIP: Contrastive language-image pretraining
CNN: Convolutional neural network
eFST: Estimated Fitzpatrick skin type
eMST: Estimated Monk Skin Tone
IQR: Interquartile range
IRB: Institutional Review Board
MIL: Multiple instance learning
OOD: Out-of-distribution
PR-AUC: Precision-recall area under the curve
ROC-AUC: Receiver operating characteristic area under the curve
SCIN: Skin Condition Image Network
ViT: Vision transformer

## Appendix A. Exploratory closed multimodal assistant comparison

This analysis was exploratory and was not a primary endpoint. It is included to contextualize performance against general-purpose multimodal assistants but should not be interpreted as a definitive benchmark because results may vary by model version, prompt design, access date, and output parsing rules.

### Appendix A.1. Zero-Shot Evaluation of Closed Multimodal Models

To contextualize the performance of DermAssist, we conducted a comparative evaluation of several closed, proprietary multimodal assistants on the same held-out test set (Figure A.6). For each case, all associated images (one to three) were provided as input to the target model’s API using a structured, zero-shot prompting strategy. The prompt instructed the model to act as a dermatological assistant and return a valid JSON object. This object was required to contain a confidence score (between 0 and 1) for each of the ten target diagnostic classes, with the explicit constraint that the scores must sum to one. A brief textual rationale was also requested under a separate ‘notes’ key.

Model responses were programmatically parsed to extract the 10-dimensional probability vector for each case; any labels omitted from a model’s response were assigned a probability of zero. These vectors were then used to compute the same performance metrics (e.g., ROC-AUC) as our primary models, enabling a direct comparison. No in-context examples or exemplars were provided, ensuring a strict zero-shot evaluation protocol. The exact prompt for Gemini models was as follows:

##### Gemini-Series Prompt

You are assisting with a dermatology classification study. Each request will include multiple clinical skin images from a single patient case. The target diagnostic vocabulary contains exactly these 10 labels: Eczema, Allergic Contact Dermatitis, Insect Bite, Urticaria, Psoriasis, Folliculitis, Irritant Contact Dermatitis, Tinea, Herpes Zoster, Drug Rash. Return a strict JSON object with keys for each label and numeric confidence scores between 0 and 1 inclusive. Scores should sum to 1 across the 10 labels. Include an additional key ‘notes’ with brief reasoning. Do not include markdown, plain text, or extra commentary—only JSON.

**Figure A.6:**
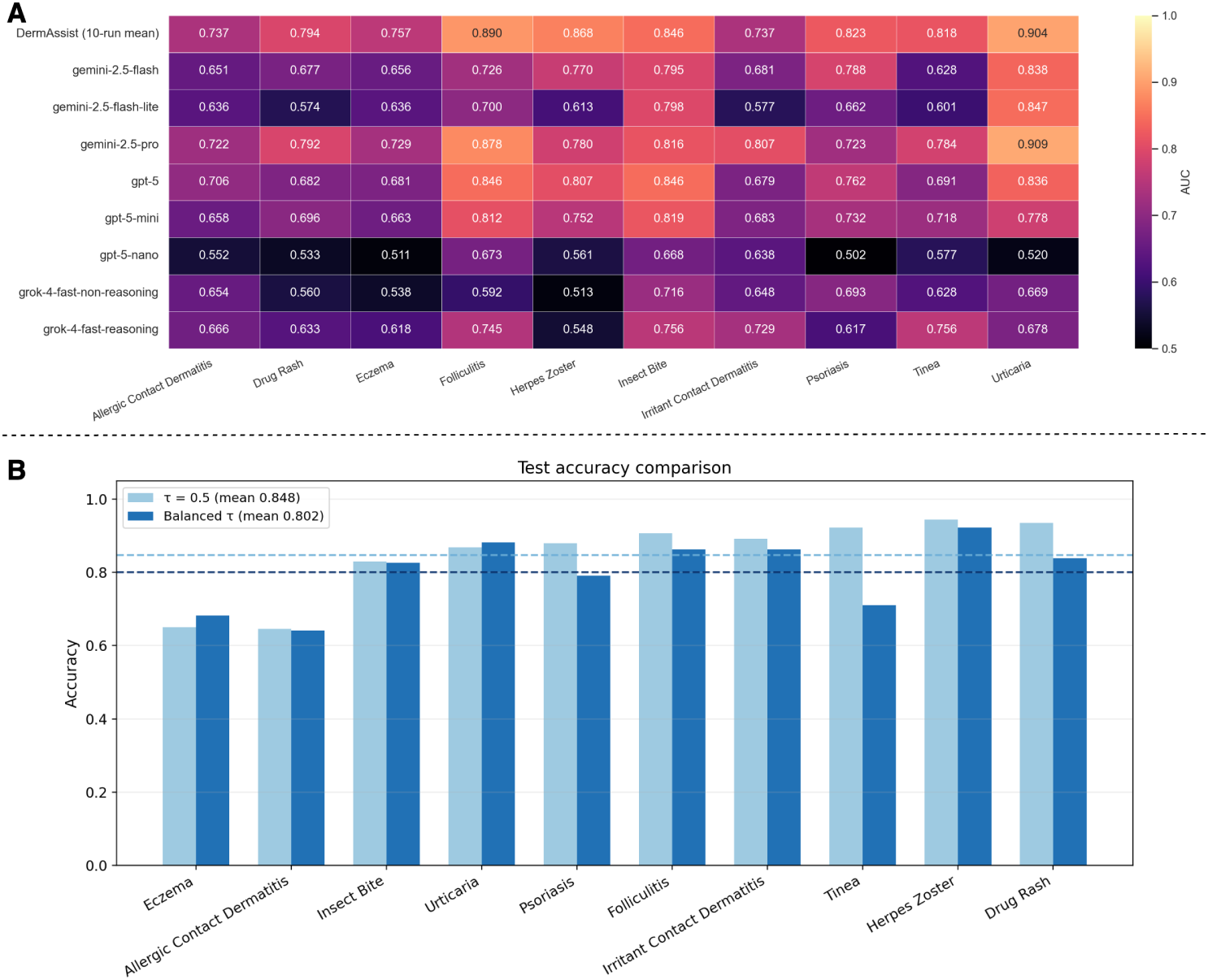
Exploratory closed multimodal assistant comparison. **(A)** Heatmap comparing the per-condition test AUCs across DermAssist and several closed multimodal assistants queried with the same multi-image cases. These exploratory results are supplementary because model versions, prompting behavior, access date, and output parsing rules may affect performance.

## Appendix B. Supplementary training diagnostics

**Figure B.7:**
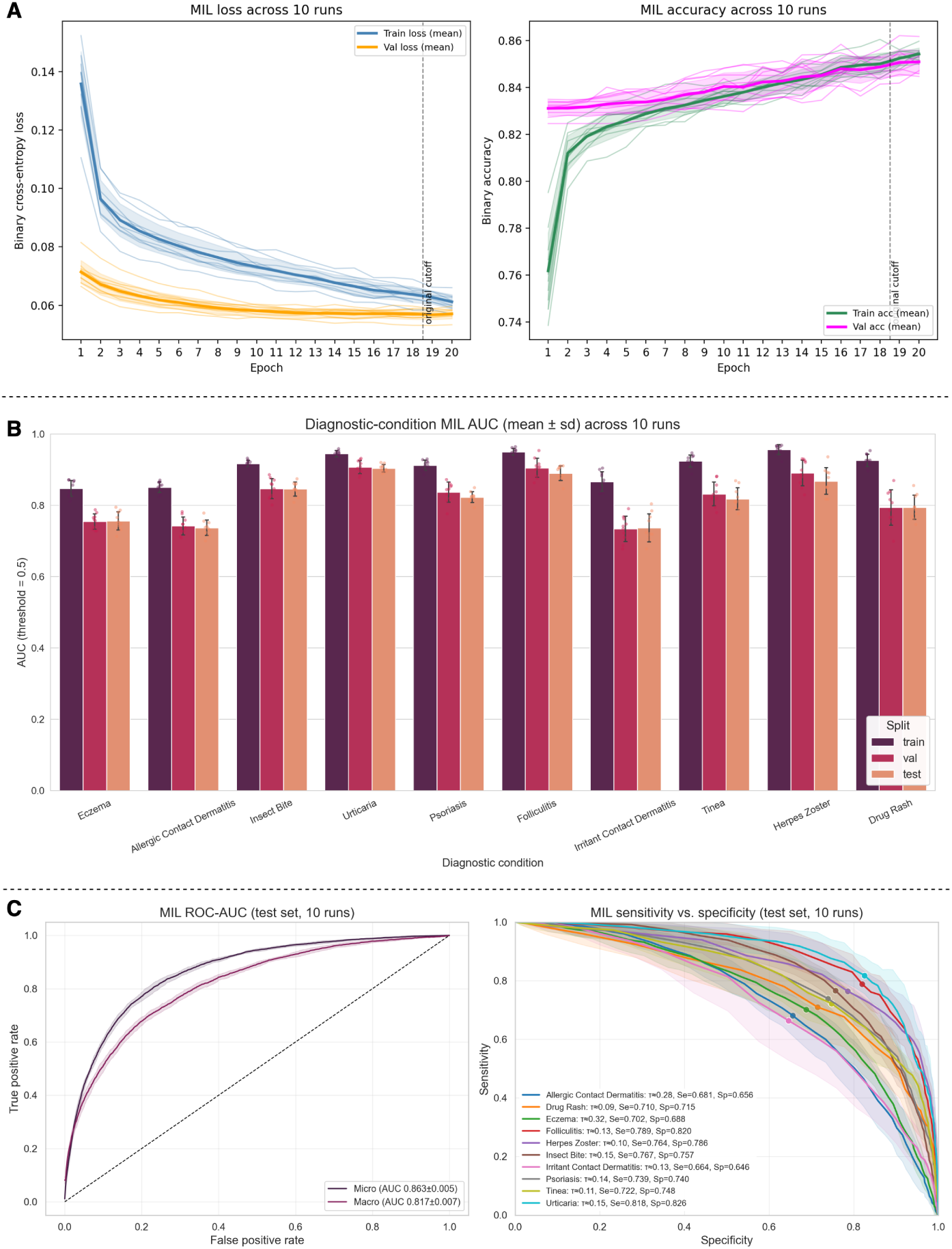
Supplementary training diagnostics for DermMIL. **(A)** Training curves for loss and accuracy across 10 runs show convergence behavior and validation stability. **(B)** Bar plots summarize per-condition AUC across training, validation, and test sets. **(C)** Aggregated micro and macro ROC curves and sensitivity–specificity curves provide additional model diagnostics.

## Appendix C. Dataset Overview

The public SCIN source resource contains 5,033 filtered contributions and approximately 10,408 images. The DermAssist analytic cohort used for modeling is a 10-condition subset comprising 2,336 cases and 5,041 image rows/embeddings. Table C.5 lists the 30 most common case labels, while Figure C.8 visualises overall label balance. Each encounter contains one to three photos (mean 2.1); the distribution of bag sizes and photographic styles is shown in Figure C.9.

Key dataset statistics:

- Public SCIN source resource: 5,033 filtered contributions and approximately 10,408 images.
- DermAssist analytic cohort: 2,336 cases and 5,041 image rows/embeddings.
- DermAssist target vocabulary: eczema, allergic contact dermatitis, insect bite, urticaria, psoriasis, folliculitis, irritant contact dermatitis, tinea, herpes zoster, and drug rash.

**Table C.5:** Top 30 diagnostic labels in the DermAssist cohort.

| Case Count | count |
| --- | --- |
| Eczema | 488 |
| Allergic Contact Dermatitis | 270 |
| Urticaria | 214 |
| Insect Bite | 185 |
| Folliculitis | 142 |
| Psoriasis | 109 |
| Tinea | 93 |
| Herpes Zoster | 68 |
| Drug Rash | 58 |
| Impetigo | 40 |
| Irritant Contact Dermatitis | 39 |
| Herpes Simplex | 36 |
| Pityriasis rosea | 32 |
| Continued on next page |  |

Table C.5: Top 30 diagnostic labels in the DermAssist cohort.
| Case Count | count |
| --- | --- |
| Viral Exanthem | 24 |
| Acne | 24 |
| Keratosis pilaris | 23 |
| Lichen planus/lichenoid eruption | 22 |
| Pigmented purpuric eruption | 22 |
| Hypersensitivity | 22 |
| CD - Contact dermatitis | 21 |
| Lichen Simplex Chronicus | 21 |
| Acute dermatitis, NOS | 18 |
| Intertrigo | 17 |
| Scabies | 17 |
| Abrasion, scrape, or scab | 17 |
| Tinea Versicolor | 16 |
| Photodermatitis | 14 |
| Cellulitis | 12 |
| Granuloma annulare | 12 |
| Stasis Dermatitis | 11 |

**Figure C.8:**
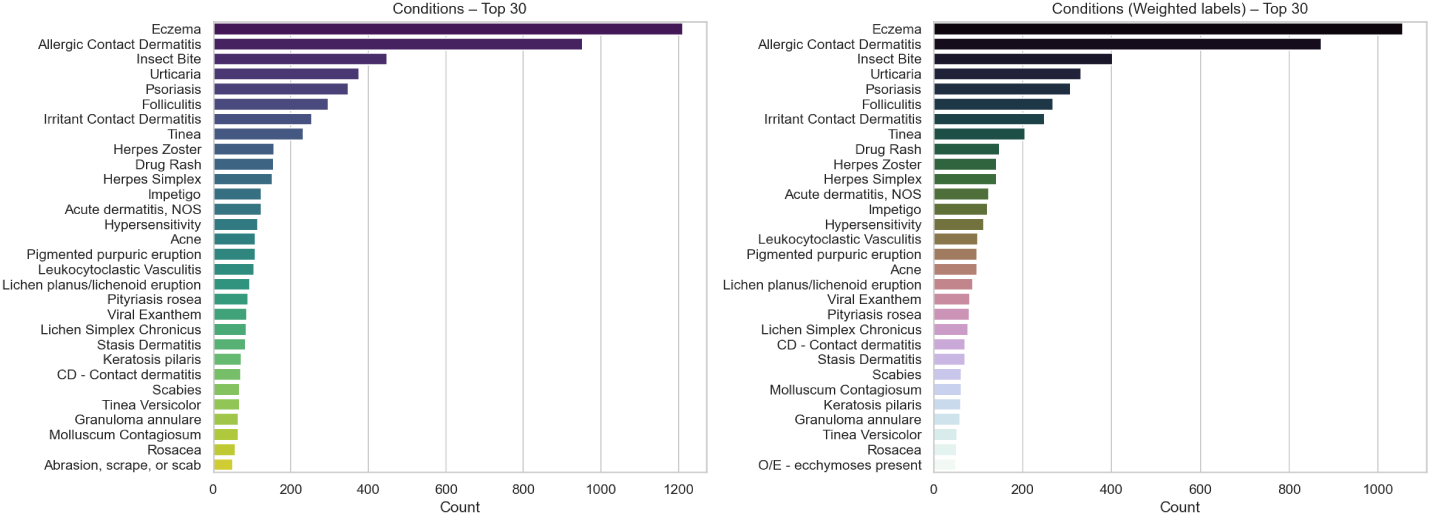
Case-level prevalence across the ten target conditions in the DermAssist cohort.

**Figure C.9:**
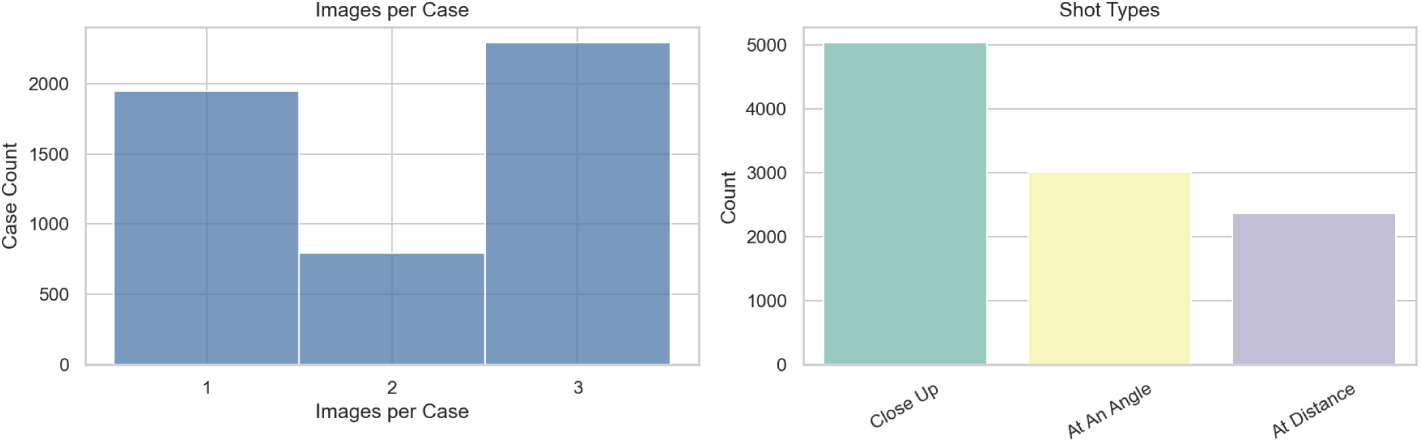
Bag sizes (left) and image framing styles (right) across the dataset.

## Appendix D. Data Quality Diagnostics

We monitor annotation quality and model supervision signals via intake confidence ratings and dermatology fellow gradability assessments. Figure D.10 contrasts clinician confidence with patient self-reported certainty, while Figure D.11 illustrates gradability scores across images. The MIL split softness plot (Figure D.12) summarises overlap between the selected ten-class subset and the broader SCIN label space.

**Figure D.10:**
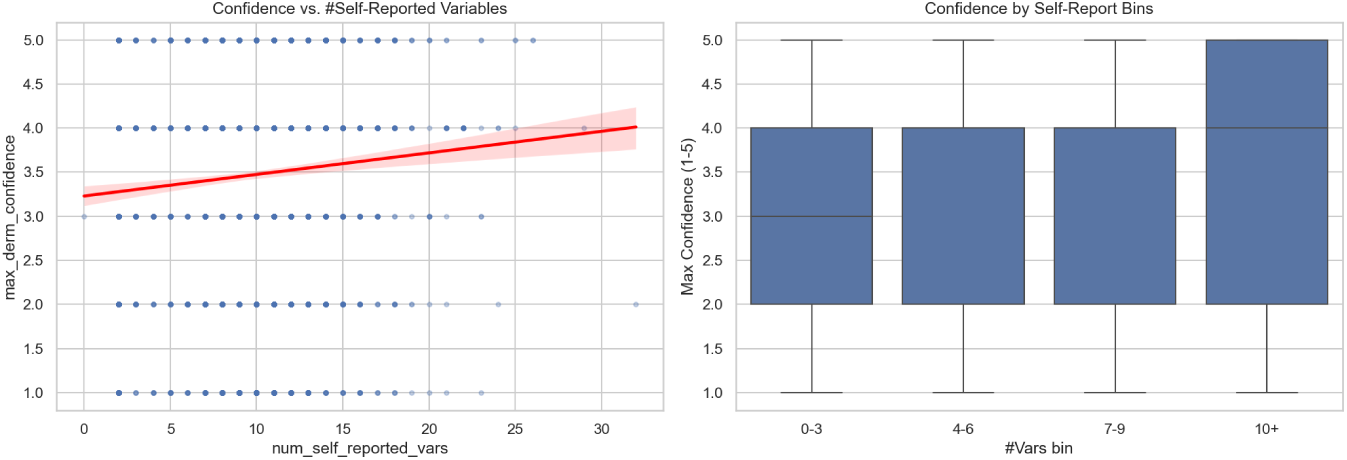
Clinician confidence ratings versus patient self-reported confidence.

**Figure D.11:**
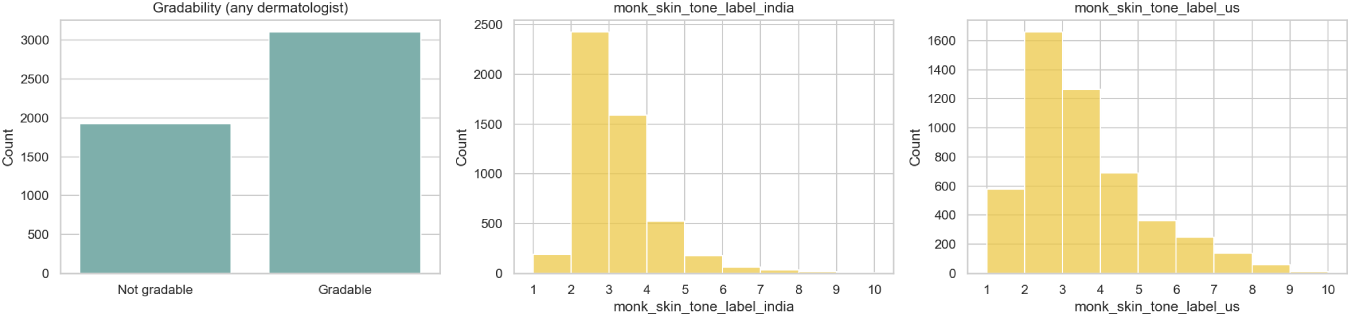
Gradability assessments indicating whether images are diagnostically usable.

**Figure D.12:**
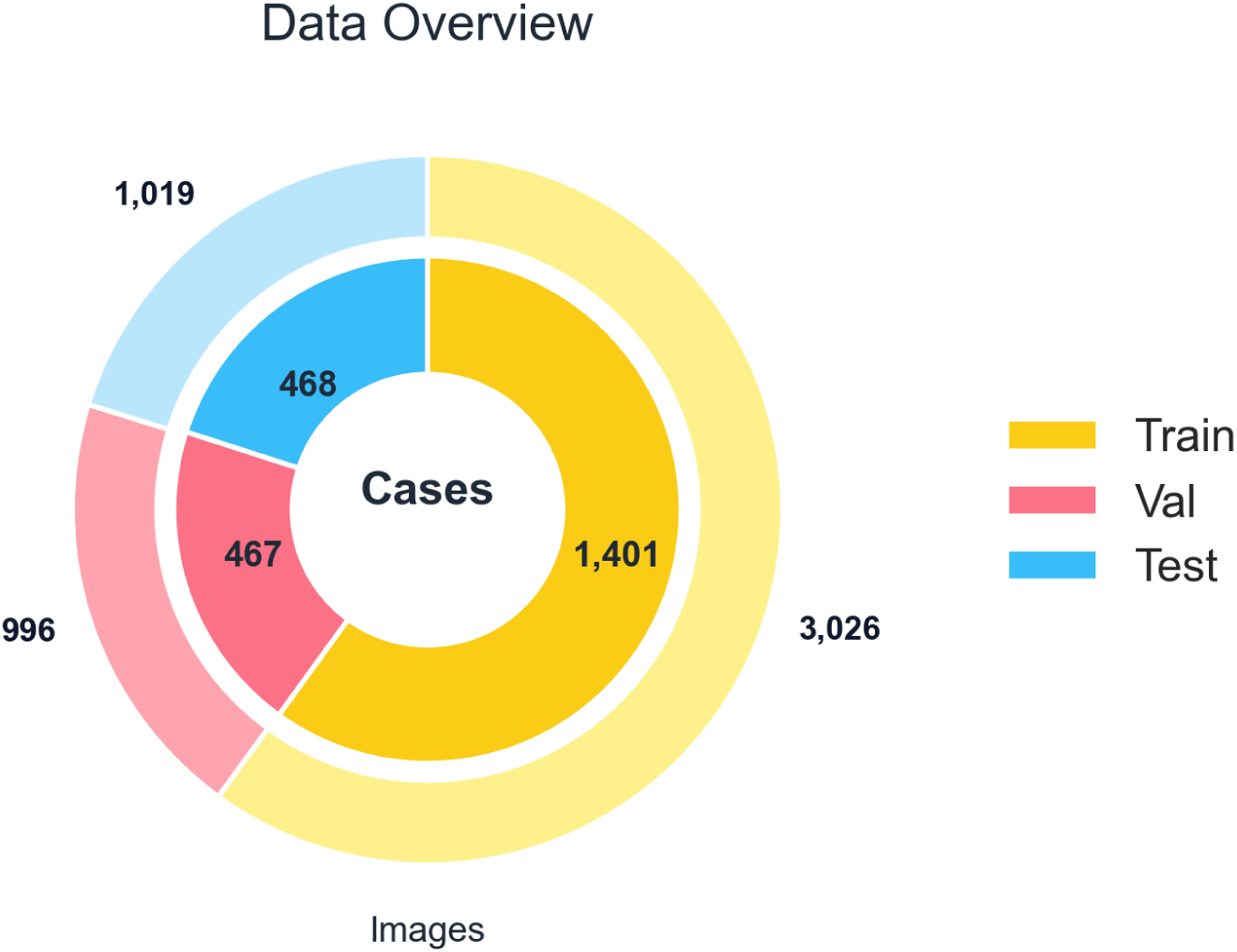
Label-set overlap heatmap between MIL training classes and the broader SCIN taxonomy (darker indicates greater co-occurrence).

## Appendix E. Hyperparameter Search Details

The Hyperband sweep (five brackets × five epochs) explored architectural depth, dropout, loss variants, and optimiser settings for the DermMIL head. Table E.6 reproduces the top trials used to select the production configuration (highlighting the cb_bce loss with no dropout, (768, 128) instance stack, and AdamW at 1.1 × 10^−4^).

**Table E.6:** Top Hyperband trials for DermMIL with Derm-Foundation embeddings. *γ* and *α* appear for focal variants; for cb_bce they are unused.

| Trial | Val ROC-AUC | Val PR-AUC | Dropout | Instance units | Attention | Bag units | Loss | $\beta$ | $\gamma$ | $\alpha$ | LR | WD | Batch |
| --- | --- | --- | --- | --- | --- | --- | --- | --- | --- | --- | --- | --- | --- |
| 1 | 0.809 | 0.493 | 0.00 | (768, 128) | 128 | (128, 64) | cb_bce | 0.996 | 2.0 | 0.00 | $1.07 \times 10^{-4}$ | $2.0 \times 10^{-6}$ | 16 |
| 0 | 0.736 | 0.398 | 0.30 | (256, 256) | 64 | (384, 128) | cb_bce | 0.953 | 2.0 | 0.00 | $2.65 \times 10^{-3}$ | $7.1 \times 10^{-5}$ | 64 |
| 4 | 0.710 | 0.362 | 0.30 | (768, 256) | 64 | (256, 128) | bce | 0.979 | 1.0 | 0.25 | $2.39 \times 10^{-3}$ | $4.31 \times 10^{-4}$ | 64 |
| 2 | 0.588 | 0.243 | 0.40 | (768, 512) | 64 | (128, 64) | cb_focal | 0.972 | 2.0 | 0.25 | $1.04 \times 10^{-3}$ | $3.0 \times 10^{-6}$ | 16 |
| 3 | 0.533 | 0.192 | 0.30 | (256, 256) | 256 | (128, 256) | bce | 0.970 | 1.0 | 0.00 | $1.6 \times 10^{-5}$ | $1.8 \times 10^{-5}$ | 64 |

## Appendix F. Class Imbalance Diagnostics

Table F.7 summarises per-class counts across splits and the resulting effective-number weights (normalised to mean one, *β* = 0.996). These weights feed directly into the CB-BCE loss described in Section 2.7.

The class-balanced binary cross-entropy objective used effective-number weights computed from the training split only. Let *n_k_* be the number of positive training cases for class *k*. Following Cui *et al.*, the unnormalised class weight is

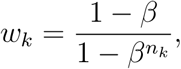

which is normalised to mean one across the *K* target classes:

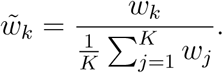

For a binary label *y* ∈ {0, 1} and predicted probability *p* ∈ (0, 1), the perlabel contribution is

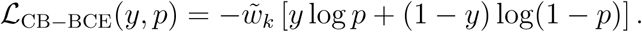

These weights increase the relative contribution of rarer positive classes while preserving a stable binary cross-entropy objective.

**Table F.7:**
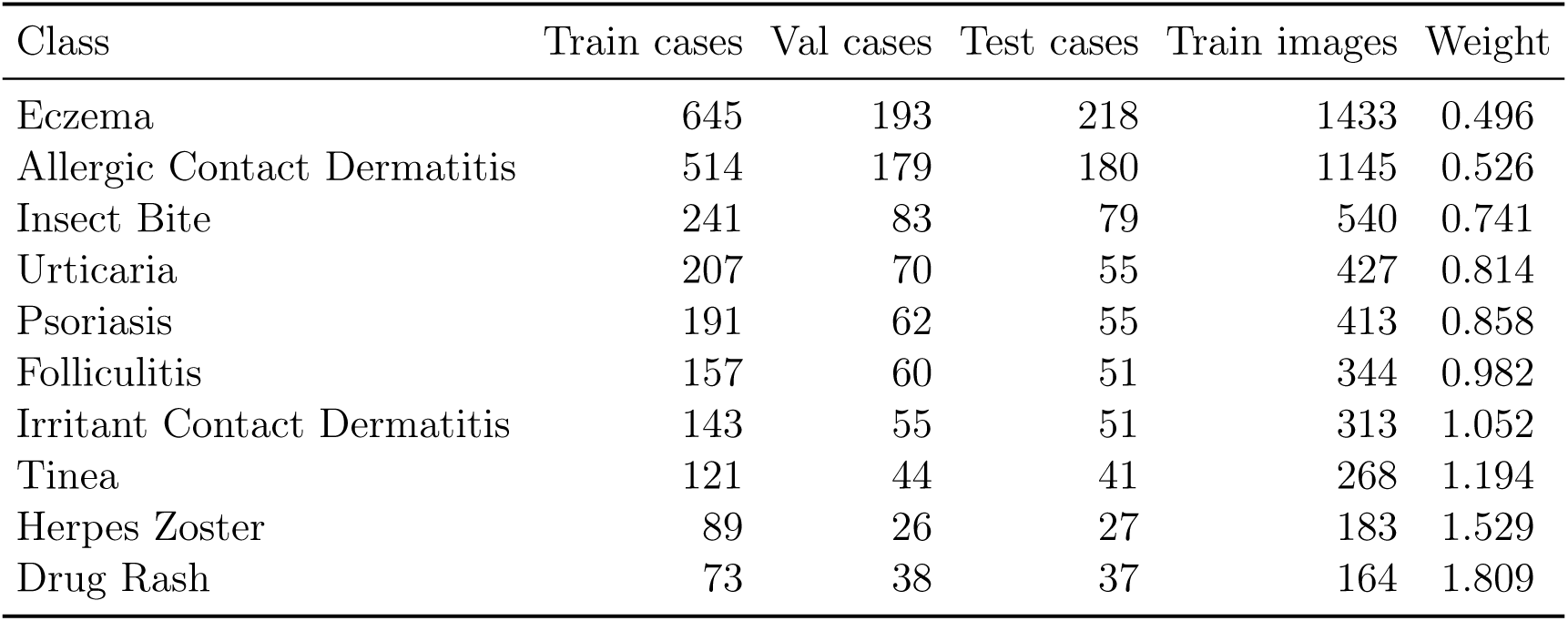
Per-class prevalence (cases) and effective-number weights. Images refer to training split counts.

| Class | Train cases | Val cases | Test cases | Train images | Weight |
| --- | --- | --- | --- | --- | --- |
| Eczema | 645 | 193 | 218 | 1433 | 0.496 |
| Allergic Contact Dermatitis | 514 | 179 | 180 | 1145 | 0.526 |
| Insect Bite | 241 | 83 | 79 | 540 | 0.741 |
| Urticaria | 207 | 70 | 55 | 427 | 0.814 |
| Psoriasis | 191 | 62 | 55 | 413 | 0.858 |
| Folliculitis | 157 | 60 | 51 | 344 | 0.982 |
| Irritant Contact Dermatitis | 143 | 55 | 51 | 313 | 1.052 |
| Tinea | 121 | 44 | 41 | 268 | 1.194 |
| Herpes Zoster | 89 | 26 | 27 | 183 | 1.529 |
| Drug Rash | 73 | 38 | 37 | 164 | 1.809 |

## Appendix G. Threshold Sweeps and Operating Points

We sweep thresholds on the validation split for each of the ten bag seeds, averaging the resulting sensitivity/specificity curves to obtain smooth mean±SD envelopes (Section 3). Table G.8 reports the conservative 0.5 base-line, while Table G.9 captures the balance-point operating points used for qualitative reporting. Supplementary Figure G.13 visualises the underlying sweeps.

**Table G.8:** Per-class test sensitivity/specificity at the default threshold *τ* = 0.5 (10-run mean).

| Class | $Se@0.5$ | $Sp@0.5$ |
| --- | --- | --- |
| Eczema | 0.250 | 0.938 |
| Allergic Contact Dermatitis | 0.080 | 0.977 |
| Insect Bite | 0.090 | 0.990 |
| Urticaria | 0.210 | 0.995 |
| Psoriasis | 0.119 | 0.995 |
| Folliculitis | 0.193 | 0.995 |
| Irritant Contact Dermatitis | 0.002 | 1.000 |
| Tinea | 0.049 | 0.999 |
| Herpes Zoster | 0.136 | 0.998 |
| Drug Rash | 0.017 | 0.999 |

**Table G.9:**
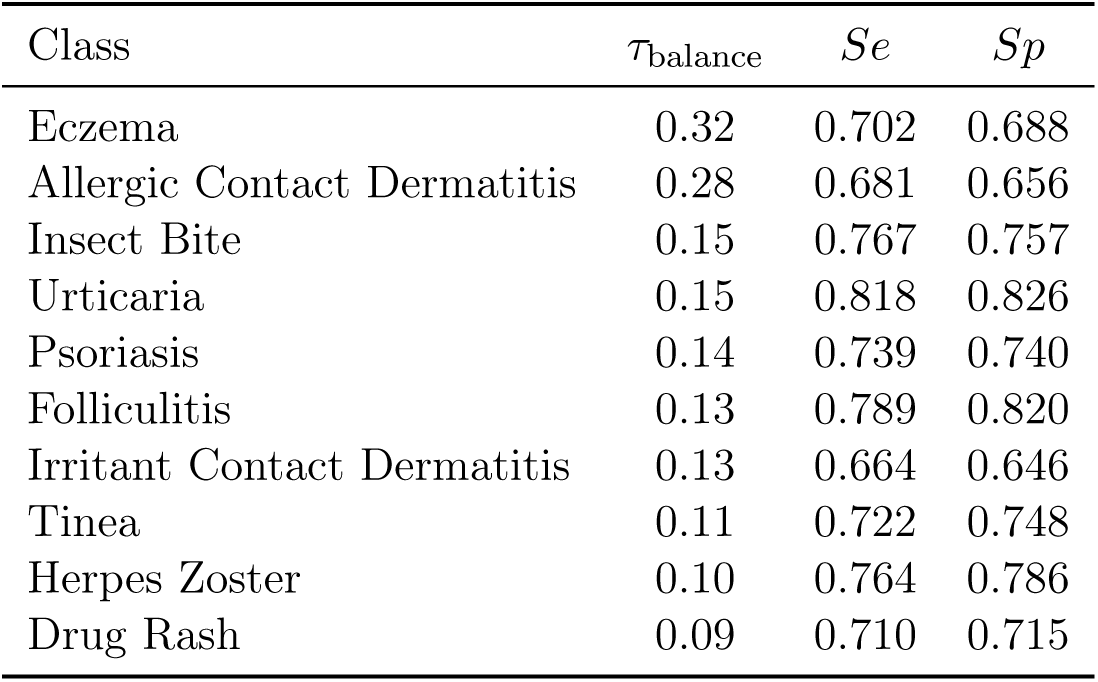
Per-class balance-point thresholds (smallest *τ* minimising |*Se* − *Sp*|) and corresponding performance (10-run mean).

**Figure G.13:**
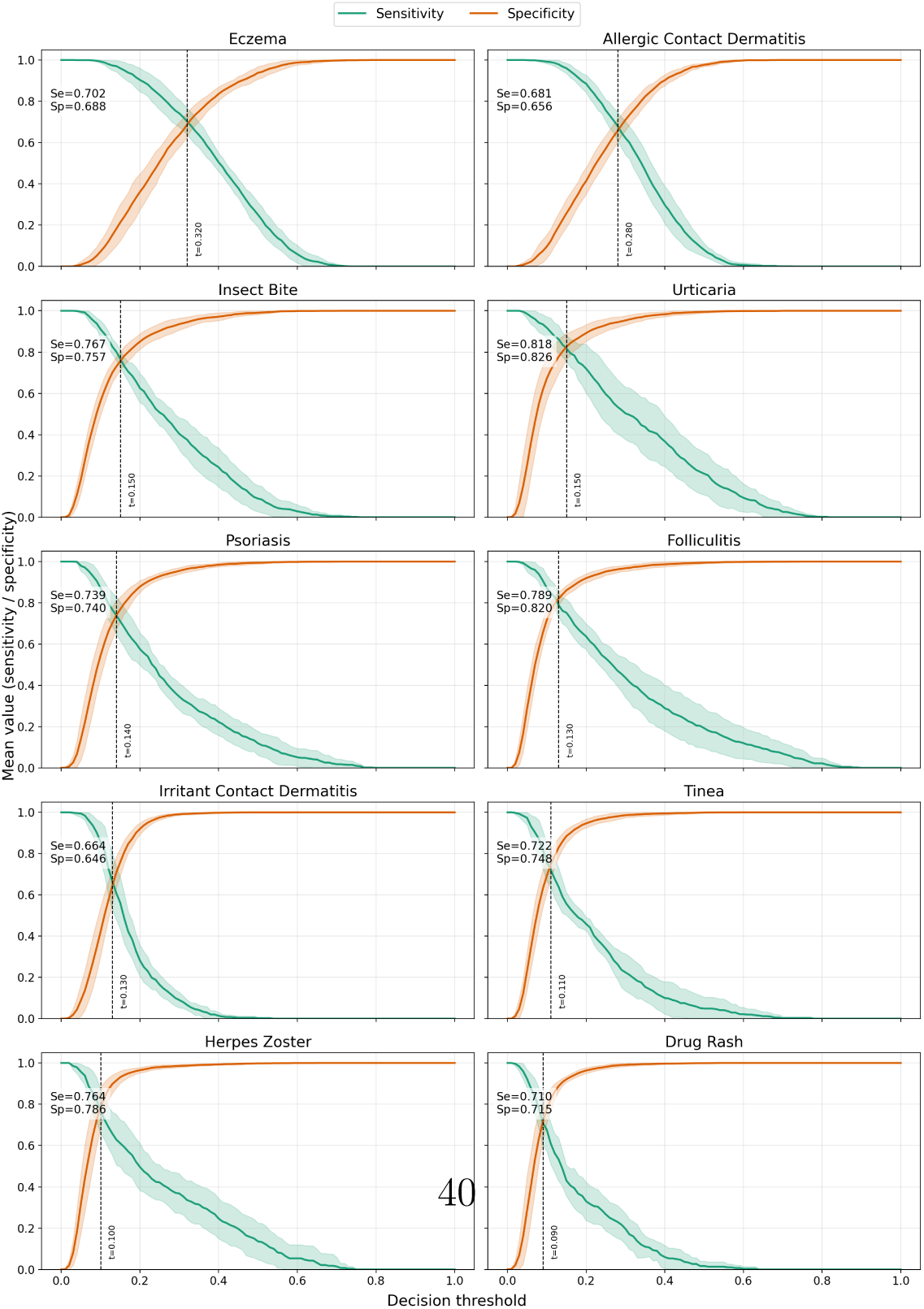
Sensitivity and specificity sweeps across thresholds for all ten diagnostic classes (mean ± SD across ten stratified bag seeds). Balance-point thresholds are shown as vertical markers.

## Appendix H. Inference Latency Benchmarks

Table H.10 summarises per-case inference latency for the DermAssist pipeline (image preprocessing, Google Derm embedding lookup, MIL head), measured on an Apple M4 workstation. Reported numbers are wall-clock seconds; entries with an asterisk are extrapolated from the eight-thread run.

**Table H.10:** Per-case end-to-end latency (seconds) by thread count.

| Batch | auto | 1 | 2 | 4 | 8 | 16* | 32* |
| --- | --- | --- | --- | --- | --- | --- | --- |
| 1 case | 6.36 | 20.65 | 12.12 | 7.89 | 6.79 | 3.42 | 1.75 |
| 3 cases | 12.17 | 45.85 | 25.84 | 17.14 | 19.19 | 9.62 | 4.87 |
| 10 cases | 85.39 | 140.91 | 94.33 | 73.32 | 56.97 | 28.59 | 14.38 |

The near-linear scaling between two and eight threads enables modest batching to amortise embedding I/O, while the single-case path remains within ∼ 6−7 seconds in auto-tuned mode.

## Appendix I. Supplementary Model Visualisations

To complement the main text comparisons, Figures I.14–I.16 present aggregate closed-model metrics, radar-style per-condition trends, and Youden-*J* threshold explorations referenced during early benchmarking.

**Figure I.14:**
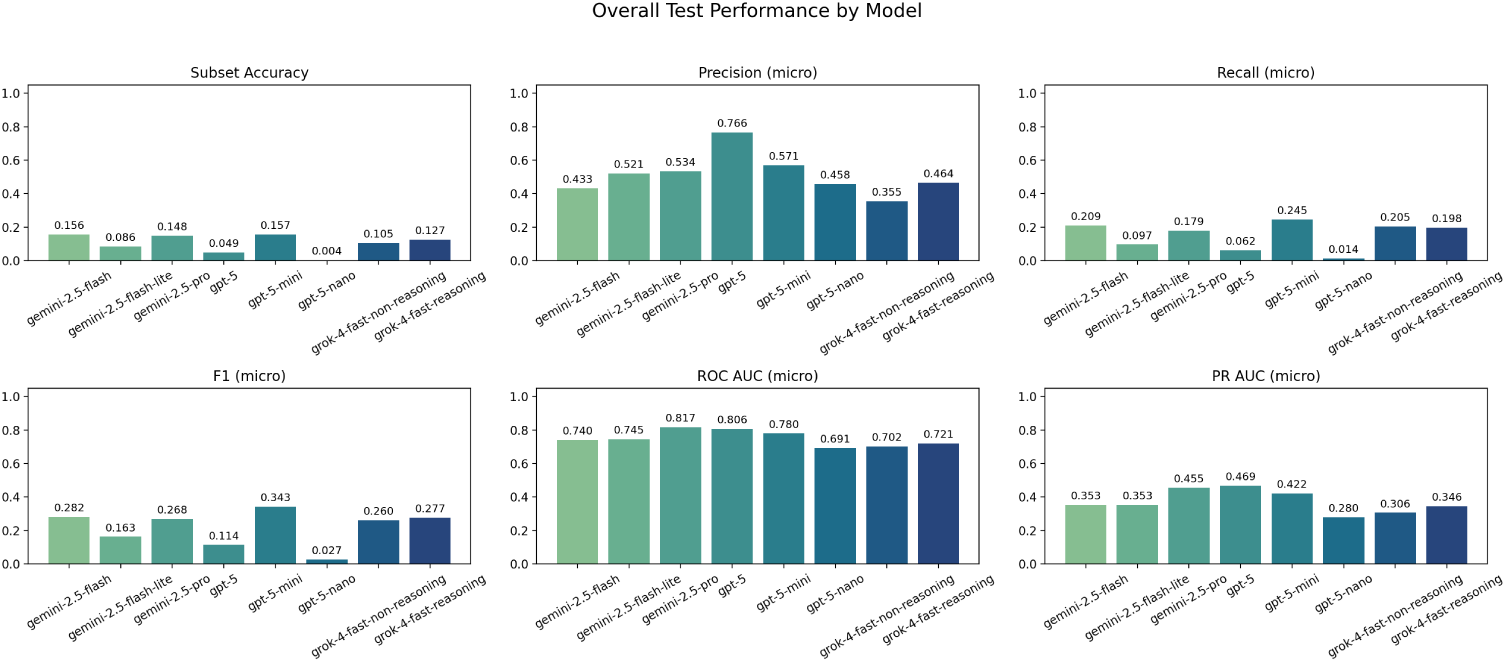
Overall test metrics for the closed-model baselines, showing accuracy, precision, recall, F1, ROC AUC, and PR AUC on the held-out split.

**Figure I.15:**
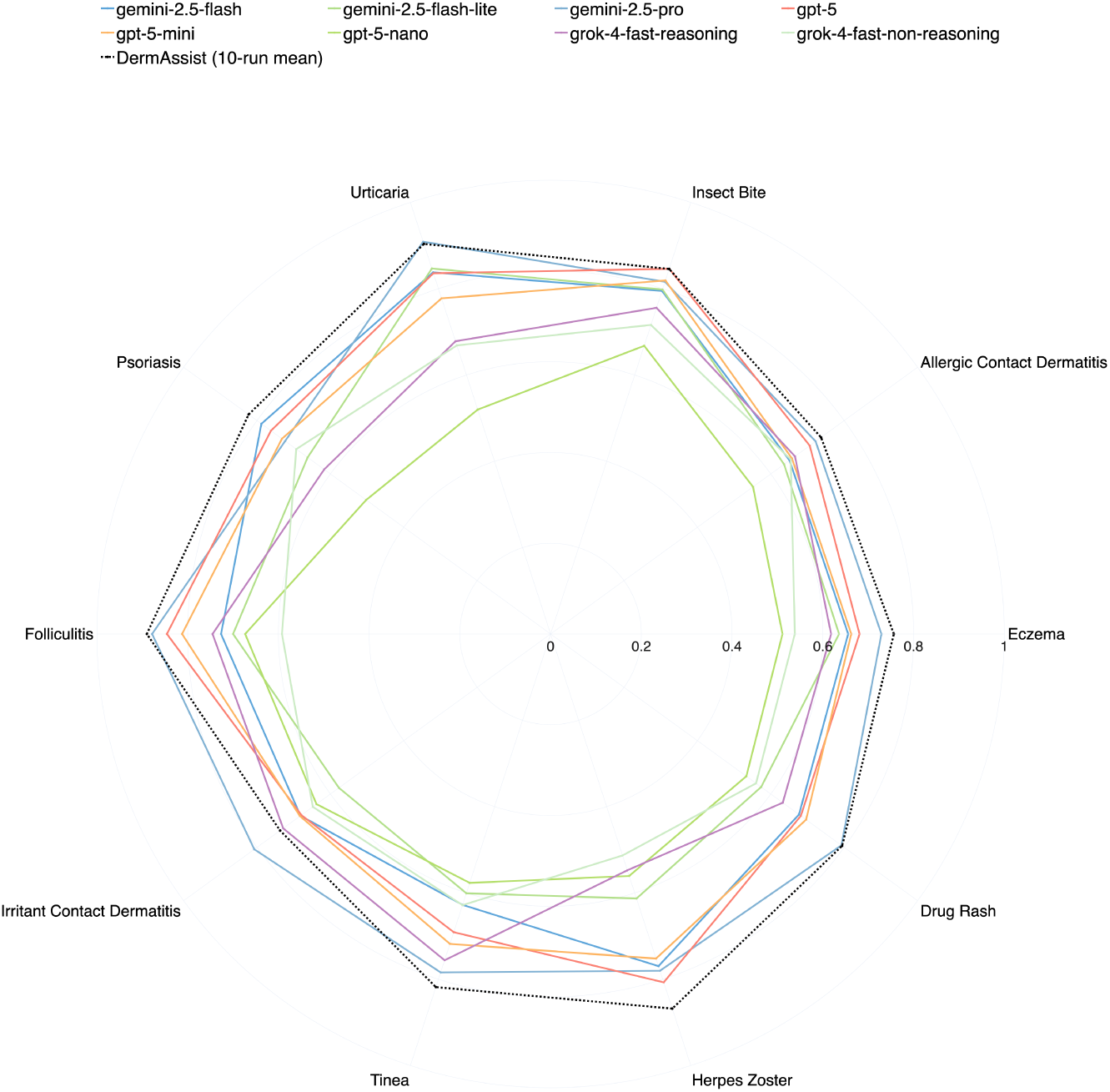
Radar comparison of per-condition ROC AUC for the closed models alongside the DermAssist 10-run mean.

**Figure I.16:**
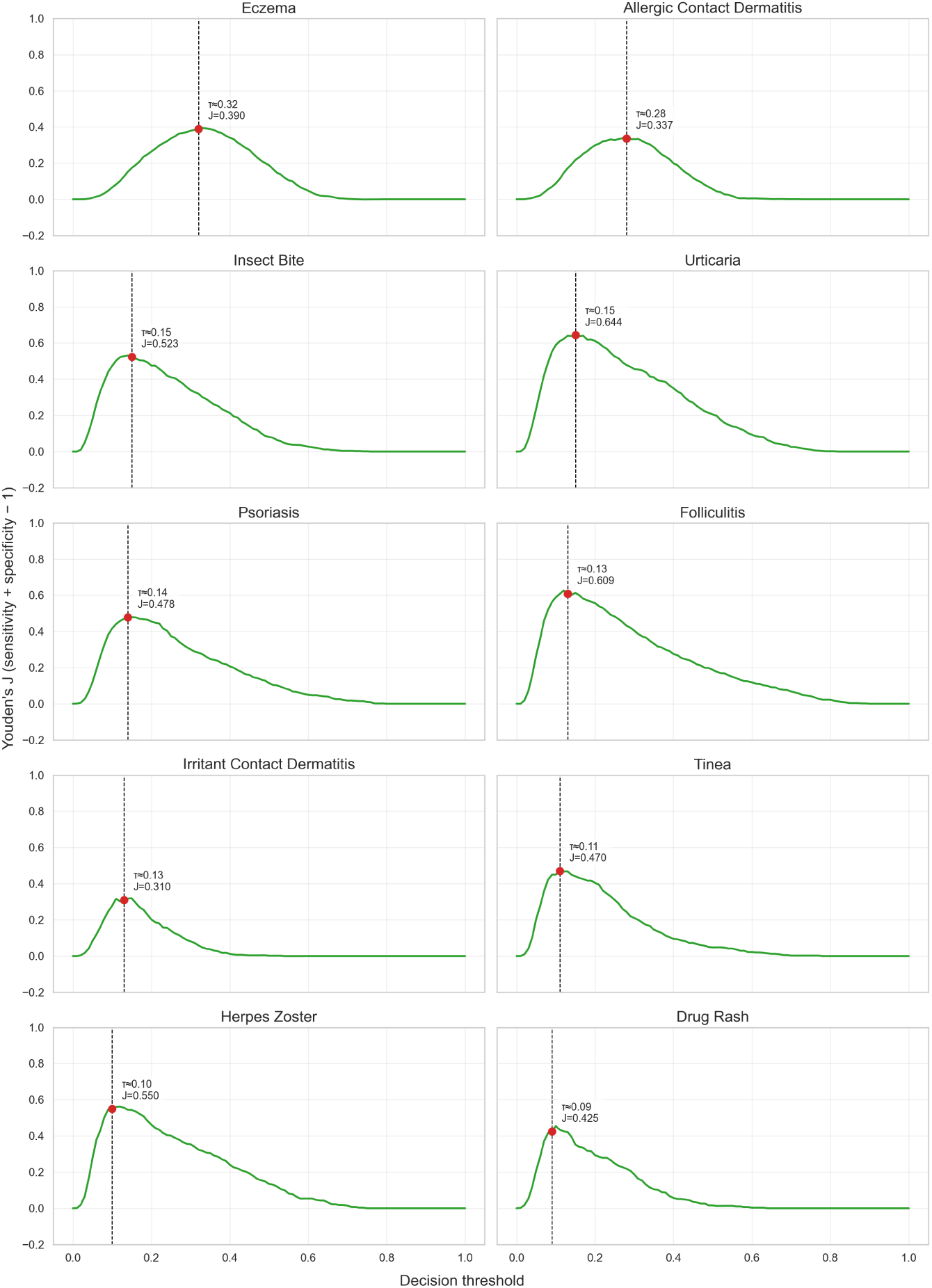
Per-condition Youden’s *J* curves (*Se* + *Sp* − 1) with optimal thresholds highlighted for DermAssist.

